# Deciphering antimicrobial resistance in bloodstream infections through clinical metagenomics

**DOI:** 10.64898/2026.04.30.26352100

**Authors:** Samruddhi Walaskar, Prajakta Jathar, Priyadarshini Mohapatra, Sindhulina Chandrasingh, Carolin Elizabeth George, Yukta Rachanwar, Rakesh Mishra, Mansi Rajendra Malik

## Abstract

**Background:** Rapid identification of pathogens and antimicrobial resistance (AMR) in bloodstream infections (BSIs) is critical for timely clinical management. Although blood culture is the reference standard, it is limited by turnaround time and incomplete resolution of resistance mechanisms. We evaluated metagenomic next-generation sequencing (mNGS) applied to flagged positive blood culture bottles to enhance diagnostic resolution and inform targeted molecular approaches.

**Methods:** Fifty-five flagged positive blood culture bottles from a tertiary care hospital in Bengaluru, India, were analyzed. Shotgun mNGS was performed directly on blood culture broth and compared with routine phenotypic identification and antimicrobial susceptibility testing (AST) from corresponding isolates. Antimicrobial resistance genes (ARGs) and plasmid replicons were profiled.

**Results:** mNGS showed high concordance with routine culture for pathogen identification (54/55; 98.2%) and improved species-level resolution across bacterial and fungal pathogens. Genotypic resistance profiles were consistent with phenotypic AST, identifying β-lactamases, efflux-associated determinants, and target modification mechanisms. Diverse ARGs and plasmid replicons (Inc-, Col-, and rep-family) were detected, providing genomic context for resistance. Sequencing predominantly reflected the cultured organism, supporting high specificity in flagged blood culture material.

**Conclusions:** mNGS applied to flagged blood culture bottles enables high-resolution characterization of pathogens and resistance determinants at a clinically actionable stage. The genomic insights generated provide a framework for developing targeted multiplex PCR assays that can reduce turnaround time and improve affordability compared with sequencing-based approaches. This strategy supports the use of mNGS as an adjunct to conventional diagnostics and as a bridge toward scalable, rapid, and cost-effective solutions for BSI diagnosis and AMR surveillance.

## Introduction

Bloodstream infections (BSIs) are major causes of morbidity and mortality and can rapidly progress to sepsis or septic shock if not diagnosed and treated promptly. Globally, BSIs and sepsis account for an estimated 48.9 million cases and 11 million deaths annually, representing nearly 20% of all deaths (1). The burden is disproportionately high in low- and middle-income countries such as India, where BSIs contribute to prolonged hospitalization, increased case-fatality rates, and significant strain on healthcare systems (2). The rising prevalence of antimicrobial resistance (AMR) further complicates management by limiting effective therapeutic options (3, 4).

The etiology of BSIs is diverse and includes both Gram-negative and Gram-positive pathogens. In India, commonly reported organisms include *Escherichia coli*, *Klebsiella pneumoniae*, *Acinetobacter baumannii*, *Pseudomonas aeruginosa*, and *Staphylococcus aureus*, many of which belong to the ESKAPEE group and are associated with healthcare-acquired infections and multidrug resistance (5). Increasing reports of carbapenem-resistant Enterobacterales and methicillin-resistant *S. aureus* underscore the need for diagnostic approaches that enable rapid pathogen identification alongside actionable resistance profiling (6).

Blood culture (BC) remains the reference standard for BSI diagnosis due to its high specificity (7, 8). However, BC-based workflows are limited by prolonged turnaround times for pathogen identification and antimicrobial susceptibility testing (AST), reduced sensitivity following prior antibiotic exposure, and incomplete recovery of fastidious or slow-growing organisms. In addition, culture-based methods provide limited insight into the genetic basis of resistance, including antimicrobial resistance genes (ARGs) and plasmid-mediated determinants that contribute to the emergence and dissemination of AMR (7, 8).

Metagenomic next-generation sequencing (mNGS) is a culture-independent approach that enables comprehensive detection of pathogens and resistance determinants directly from clinical samples (9). Previous studies have demonstrated its ability to identify fastidious organisms, detect mixed infections, and characterize ARGs and mobile genetic elements (8, 10, 11). However, sequencing from isolated colonies retains culture-associated delays, while direct sequencing from plasma is often constrained by low microbial biomass and high host DNA background, limiting diagnostic sensitivity (8, 10, 11).

Flagged positive blood culture bottles represent a clinically relevant intermediate substrate for mNGS, containing enriched microbial biomass with reduced host DNA background (12,13). Sequencing at this stage has the potential to bypass subculture-associated delays while preserving pathogen composition at the time of culture positivity, enabling high-resolution genomic characterization at a clinically actionable time point(14).

In this study, we applied mNGS directly to flagged positive blood culture bottles and compared the resulting genotypic data with routine culture identification and antimicrobial susceptibility testing (AST). We characterized pathogen profiles, antimicrobial resistance genes, and plasmid content in these samples to assess concordance with conventional diagnostics and to define the genomic landscape of bloodstream infections in this setting.

## Methods

### Study Design and Setting

This retrospective study was conducted using clinical specimens obtained in collaboration with the Department of Microbiology at Bangalore Baptist Hospital (BBH), a tertiary care center in Bengaluru, India. For metagenomic analysis, primary clinical material was obtained directly from flagged positive blood culture bottles prior to subculture. Conventional blood culture processing and routine microbiological diagnostics were performed independently as part of standard clinical care. This design enabled direct comparison between metagenomic sequencing of enriched blood culture material and hospital-generated pathogen identification and antimicrobial susceptibility testing (AST) results. The study protocol was approved by the Institutional Review Board of Bangalore Baptist Hospital.

### Blood Culture Processing and Sample Collection

Blood cultures were processed using the BACTEC™ automated blood culture system (Becton Dickinson, Sparks, MD, USA) and monitored until flagged positive. Only leftover flagged blood culture bottles with sufficient residual volume and acceptable sample integrity were included; samples not meeting these criteria were excluded. From each of the 55 eligible bottles, 250 µL of blood culture broth was aseptically collected prior to further culture manipulation. Corresponding bacterial or fungal isolates obtained through routine clinical workflows were also collected. Only samples with a single organism identified by routine culture were included in the analysis to enable paired genotypic–phenotypic comparison. Blood culture broth and isolate material were resuspended in sterile 1× phosphate-buffered saline (PBS) for downstream nucleic acid extraction and sequencing.

### Conventional Microbiology and Antimicrobial Susceptibility Testing

Microbial identification and AST were performed as part of routine diagnostics at BBH. Isolates were identified using standard phenotypic methods and tested against a panel of 28 antibiotics. Susceptibility testing was performed using the disk diffusion method, and results were interpreted according to Clinical and Laboratory Standards Institute (CLSI) guidelines (15, 16).

### Nucleic Acid Extraction

Genomic DNA was extracted from blood culture broth using the QIAamp DNA Blood Mini Kit (QIAGEN GmbH, Hilden, Germany) according to the manufacturer’s instructions. DNA concentration and quality were assessed using a Qubit™ 4 Fluorometer (Thermo Fisher Scientific Inc., Waltham, MA, USA). Extracted DNA was used for sequencing library preparation.

### Metagenomic Next-Generation Sequencing (mNGS)

Sequencing libraries were prepared from all samples <u>(55 blood culture bottles</u> using the Nextera XT DNA Library Preparation Kit (Illumina, Inc., San Diego, CA, USA; Cat. No. FC-131-1096) according to the manufacturer’s protocol. Library concentration and quality were assessed using a Qubit™ Fluorometer. Whole-genome metagenomic sequencing was performed on the Illumina NovaSeq 6000 platform, generating paired-end reads (2 × 100 bp).

### Bioinformatics Analysis

Raw sequencing data were processed using an in-house standardized mNGS pipeline. Approximately 20–80 million paired-end reads were generated per sample. Quality assessment was performed using FastQC (v0.12.1) (17), with summary reports generated using MultiQC (v1.13) (18). Adapter trimming and quality filtering were performed using Cutadapt (v1.18), and reads shorter than 35 bp were excluded (19).

Host-derived reads were removed by mapping to the human reference genome GRCh38 using the Burrows–Wheeler Aligner (BWA), retaining only unmapped reads for downstream analysis (20). Following host depletion, approximately 40–85% of reads were removed, consistent with expectations for blood culture–derived material.

Taxonomic classification was performed using Kraken2 (v2.1.3) against the maxikraken2_1903_140GB database (21). Assignments were filtered based on confidence scores and genome coverage.

Antimicrobial resistance gene (ARG) profiling was performed using the Comprehensive Antibiotic Resistance Database (CARD v3.2.8). Resistance genes were identified using the Resistance Gene Identifier (RGI v6.0.3) (22), with classification based on curated detection models. High-confidence ARGs were retained using stringent coverage and alignment thresholds. Findings were corroborated using the Chan Zuckerberg Initiative ID (CZ ID) platform (23, 24).

### Statistical Analysis and Data Visualization

All analyses were conducted using R (version 4.1). Pathogen abundance and ARG distributions were visualized using heatmaps, bar plots, and Sankey diagrams. To ensure specificity, only taxa and ARGs with 100% sequence coverage were retained for downstream analysis (12).

Data preprocessing and transformation were performed using the tidyverse framework. Visualization was implemented using ggplot2, dplyr, tidyr, and ComplexHeatmap (25). Color schemes were managed using RColorBrewer. Sankey and alluvial plots were generated using compatible flow visualization libraries (26).

## Results

### Taxonomic Distribution of Pathogens in Bloodstream Infections

Species-level taxonomic profiles of bacterial and fungal pathogens were generated from flagged positive blood culture samples using a metagenomic next-generation sequencing (mNGS) workflow incorporating Kraken2 and Chan Zuckerberg ID (CZ ID) for classification (Fig. 2). Taxonomic assignments were concordant between both analytical pipelines, supporting consistency of species-level detection. Sample-wise relative abundance data are provided in Supplementary File S1.

**Fig 1:**
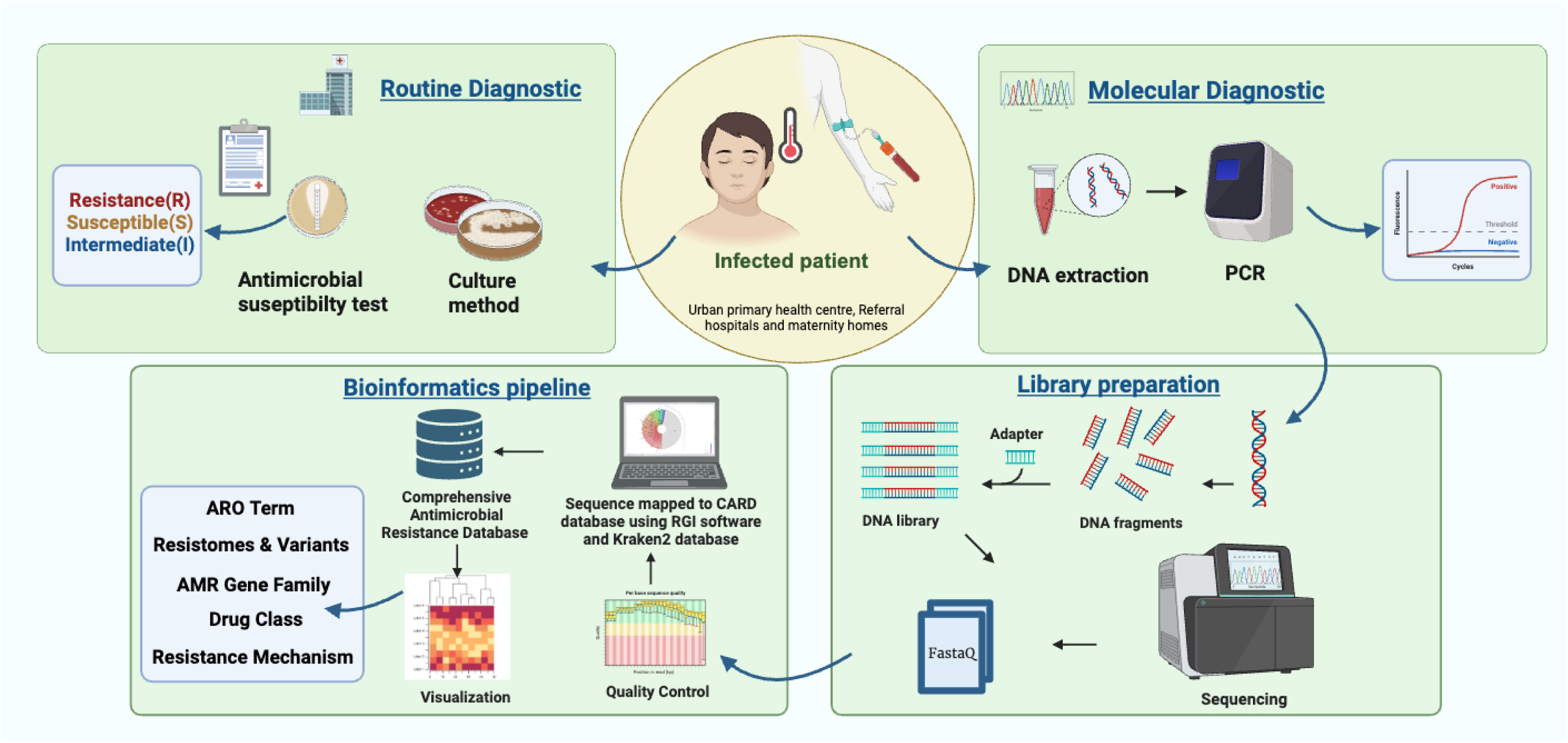
Graphical abstract outlining the experimental design and analytical workflow employed in this study.

**Fig 2.**
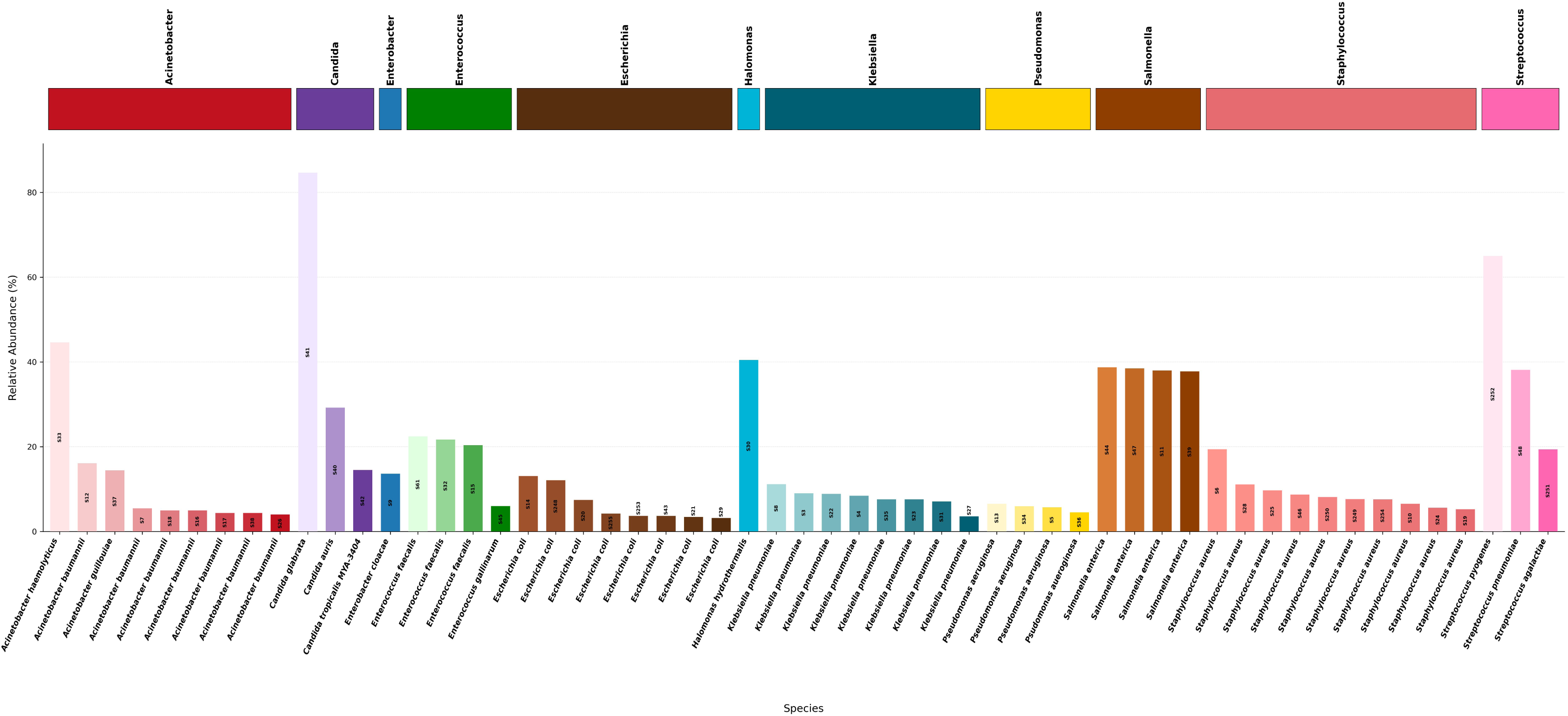
Relative abundance of bacterial and fungal species identified in positive blood culture samples (n = 55) using mNGS.

Across the 55 samples, mNGS identified a diverse range of Gram-negative and Gram-positive pathogens. *Escherichia coli* and *Klebsiella* spp. were each detected in 14.5% (8/55) of samples, while *Acinetobacter* spp. were identified in 16.4% (9/55), predominantly *Acinetobacter baumannii*. *Enterococcus* spp. were detected in 7.2% (4/55) of samples, primarily *Enterococcus faecalis*, and *Staphylococcus aureus* was identified across multiple samples at clinically relevant abundances.

Pathogen identification by mNGS showed high concordance with routine blood culture–based identification (54/55; 98.2%) (Table 1). All samples included in the analysis yielded a single organism on routine culture, enabling direct comparison of pathogen identification and resistance profiles between mNGS and conventional methods. In addition, mNGS provided species-level resolution consistent with culture-based results.

**Table 1:**
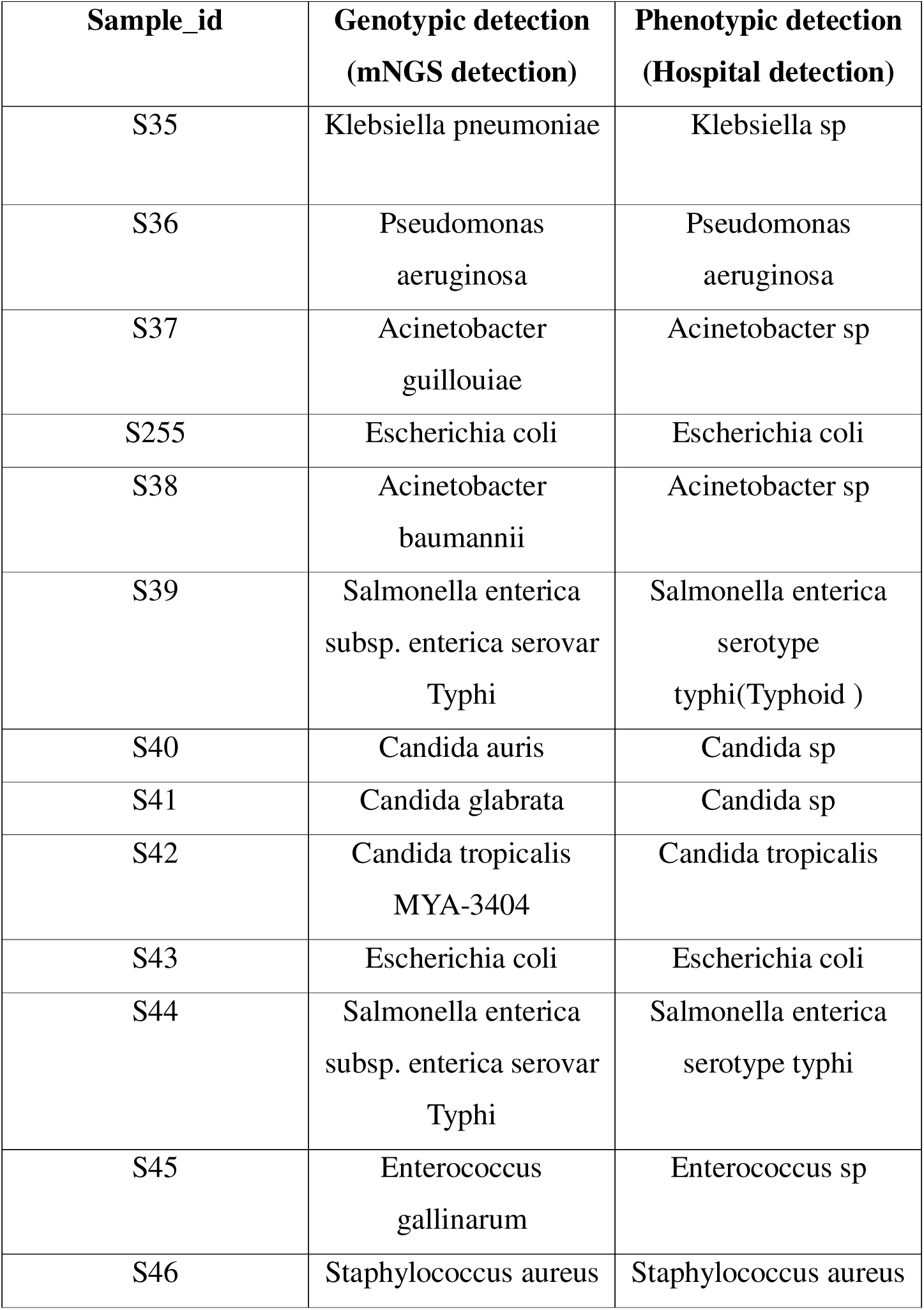
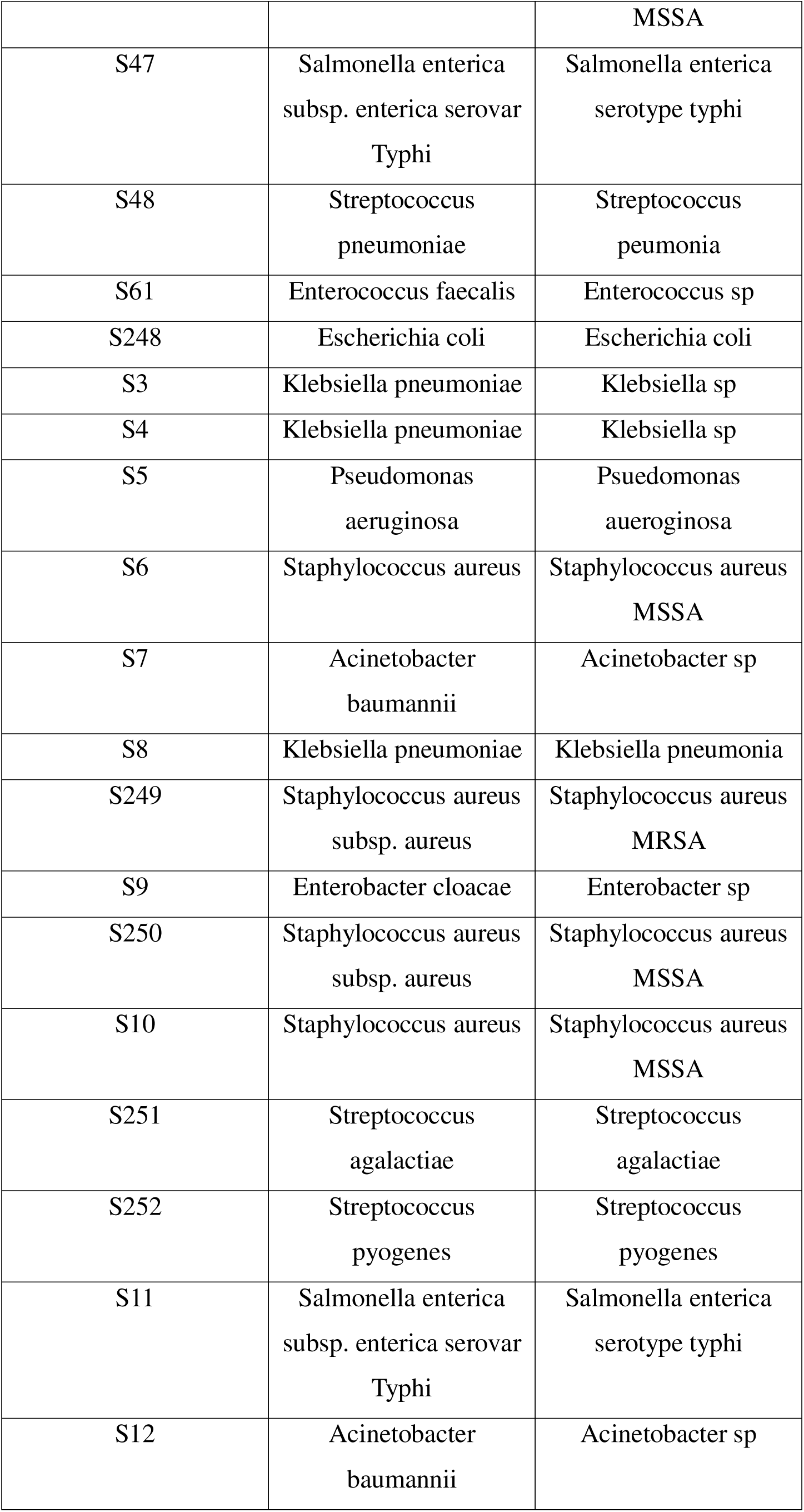
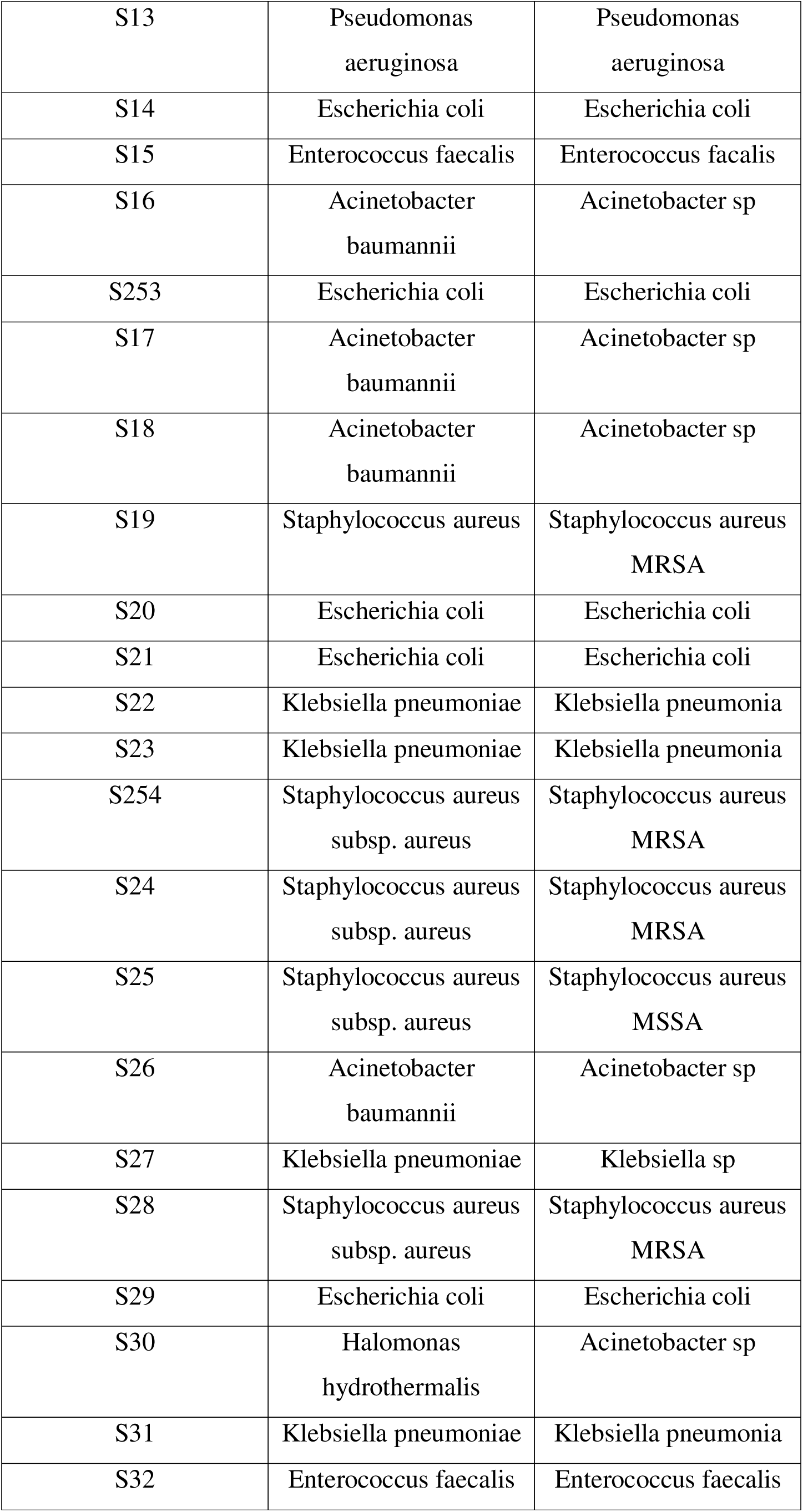
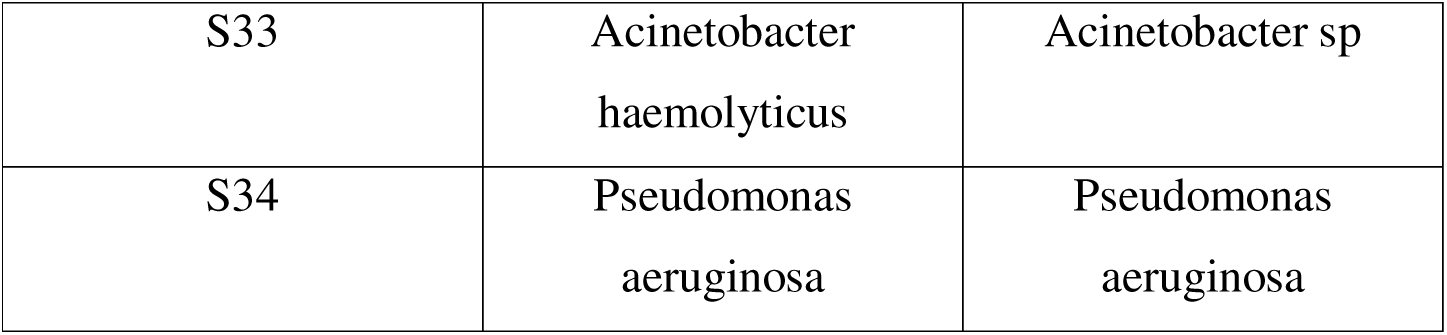
Genotypic and phenotypic detection of pathogens from flagged blood bottle samples and corresponding isolates.

Although mNGS enables detection of polymicrobial signals, sequencing profiles predominantly reflected the organism identified by routine culture. A low-abundance taxon (*Halomonas* spp.) was detected in one sample.

### Mapping Phenotypic and Genotypic Antimicrobial Resistance Across Bloodstream Infection Pathogens

Comparative analysis of antimicrobial resistance (AMR) was performed by integrating phenotypic antimicrobial susceptibility testing (AST) with metagenomic resistance gene profiles (Fig. 3). Resistance determinants were identified using the Comprehensive Antibiotic Resistance Database (CARD) and cross-validated with Chan Zuckerberg ID (CZ ID). Phenotypic AST results (Fig. 3A) and corresponding genotypic resistance profiles (Fig. 3B) are summarized across all samples, with the complete resistome provided in Supplementary File S2.

**Figure 3A.**
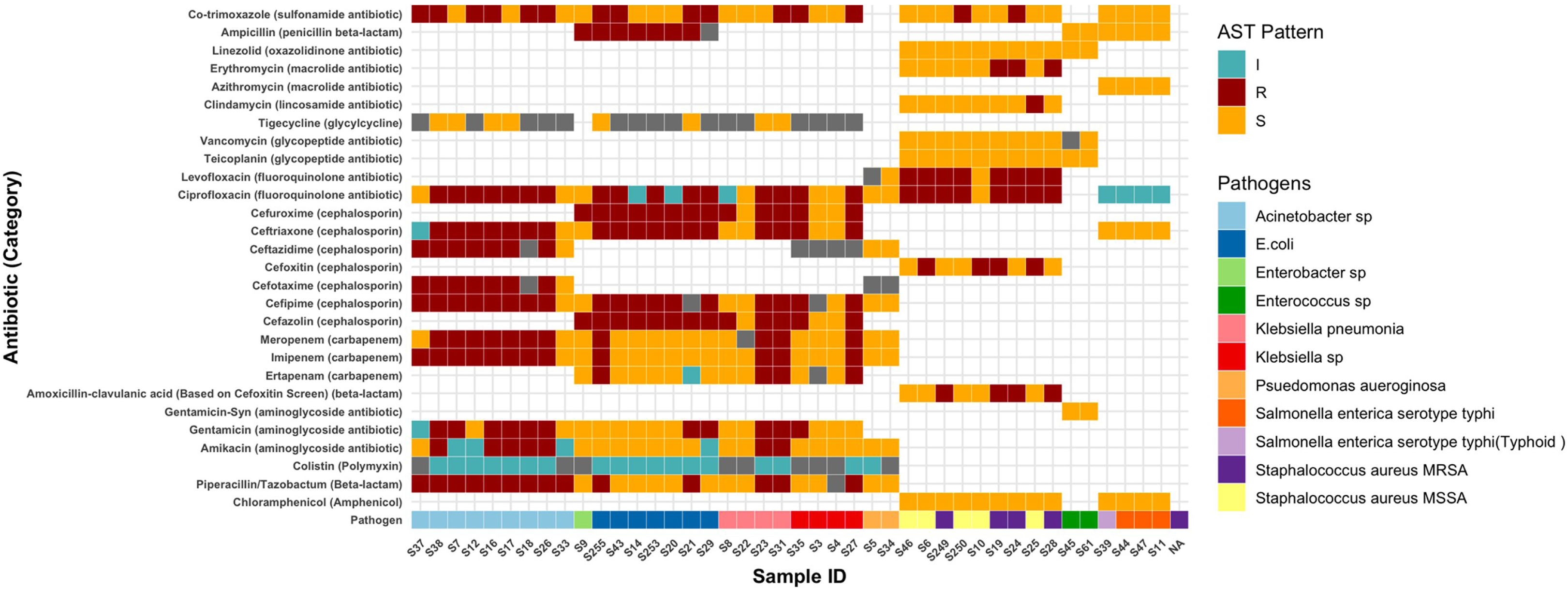
Phenotypic antimicrobial susceptibility profiles across bloodstream infection isolates.

**Figure 3B.**
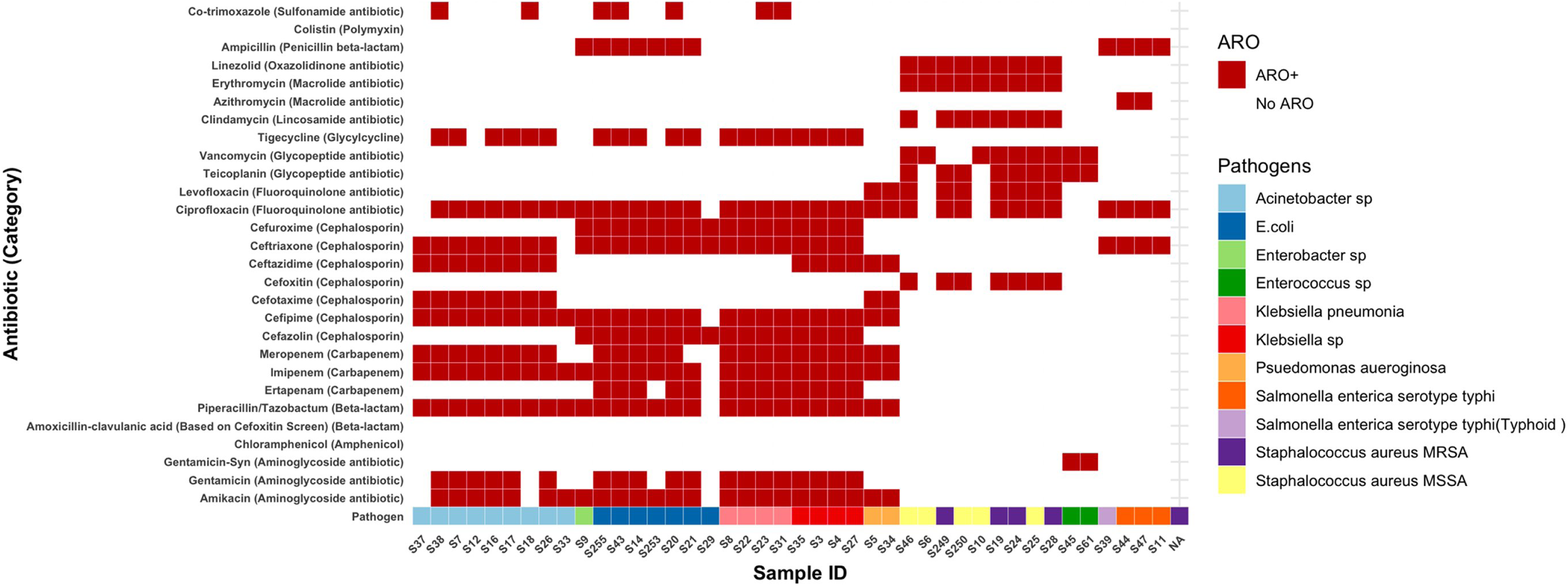
Metagenomic antimicrobial resistance gene (ARG) profiles across bloodstream infection isolates.

Among Gram-negative pathogens (*Acinetobacter* spp., *Klebsiella* spp., *Escherichia coli*, *Enterobacter* spp., *Pseudomonas aeruginosa*, and *Salmonella enterica* serovar Typhi), phenotypic resistance was most frequently observed to third- and fourth-generation cephalosporins (cefepime, cefotaxime, ceftazidime, ceftriaxone), aminoglycosides (amikacin, gentamicin), and fluoroquinolones (ciprofloxacin, levofloxacin). Reduced susceptibility to carbapenems (imipenem, meropenem) and colistin was observed in a subset of isolates, particularly *Acinetobacter* spp. and *Klebsiella pneumoniae*. Susceptibility to tigecycline, piperacillin–tazobactam, and carbapenems was retained in subsets of *E. coli*, *P. aeruginosa*, and *S. Typhi* isolates.

Genotypic analysis of Gram-negative isolates identified β-lactamases (including OXA-type and metallo-β-lactamases such as NDM-1 and VIM-2), aminoglycoside-modifying enzymes, quinolone resistance–associated targets (*gyrA*, *parC*), and multidrug efflux systems. These determinants were detected in isolates exhibiting phenotypic resistance or intermediate susceptibility, whereas phenotypically susceptible isolates showed limited or intrinsic resistance-associated genes.

Among Gram-positive pathogens (*Staphylococcus aureus*, *Enterococcus* spp., *Streptococcus agalactiae*, *Streptococcus pyogenes*, and *Streptococcus pneumoniae*), phenotypic AST demonstrated variable susceptibility profiles. Resistance to β-lactams (penicillin/oxacillin) was observed in methicillin-resistant *S. aureus* (MRSA), while macrolide and fluoroquinolone resistance was detected in a subset of isolates. Susceptibility to glycopeptides (vancomycin, teicoplanin) and linezolid was retained across isolates. Correspondingly, genotypic analysis identified methicillin resistance determinants in MRSA and macrolide- and fluoroquinolone-associated resistance markers in isolates with reduced susceptibility.

For fungal pathogens (*Candida* spp.), phenotypic testing demonstrated variable resistance to azoles (fluconazole, voriconazole), while susceptibility to echinocandins (caspofungin, micafungin) and amphotericin B was retained. Genotypic analysis identified azole-associated resistance markers in isolates with corresponding phenotypic resistance or intermediate susceptibility.

Overall, genotypic resistance profiles were consistent with phenotypic AST across pathogen groups, providing gene-level resolution aligned with culture-based susceptibility results.

### Comprehensive Profiling of Antimicrobial Resistance Gene Families and Resistance Mechanisms

Antimicrobial resistance (AMR) gene profiles were analyzed to characterize resistance mechanisms across bloodstream infection pathogens. Relationships between samples, identified pathogens, AMR gene families, and resistance mechanisms are visualized using a Sankey plot (Fig. 4). The complete catalogue of resistance genes and functional annotations is provided in Supplementary File S3.

**Figure 4.**
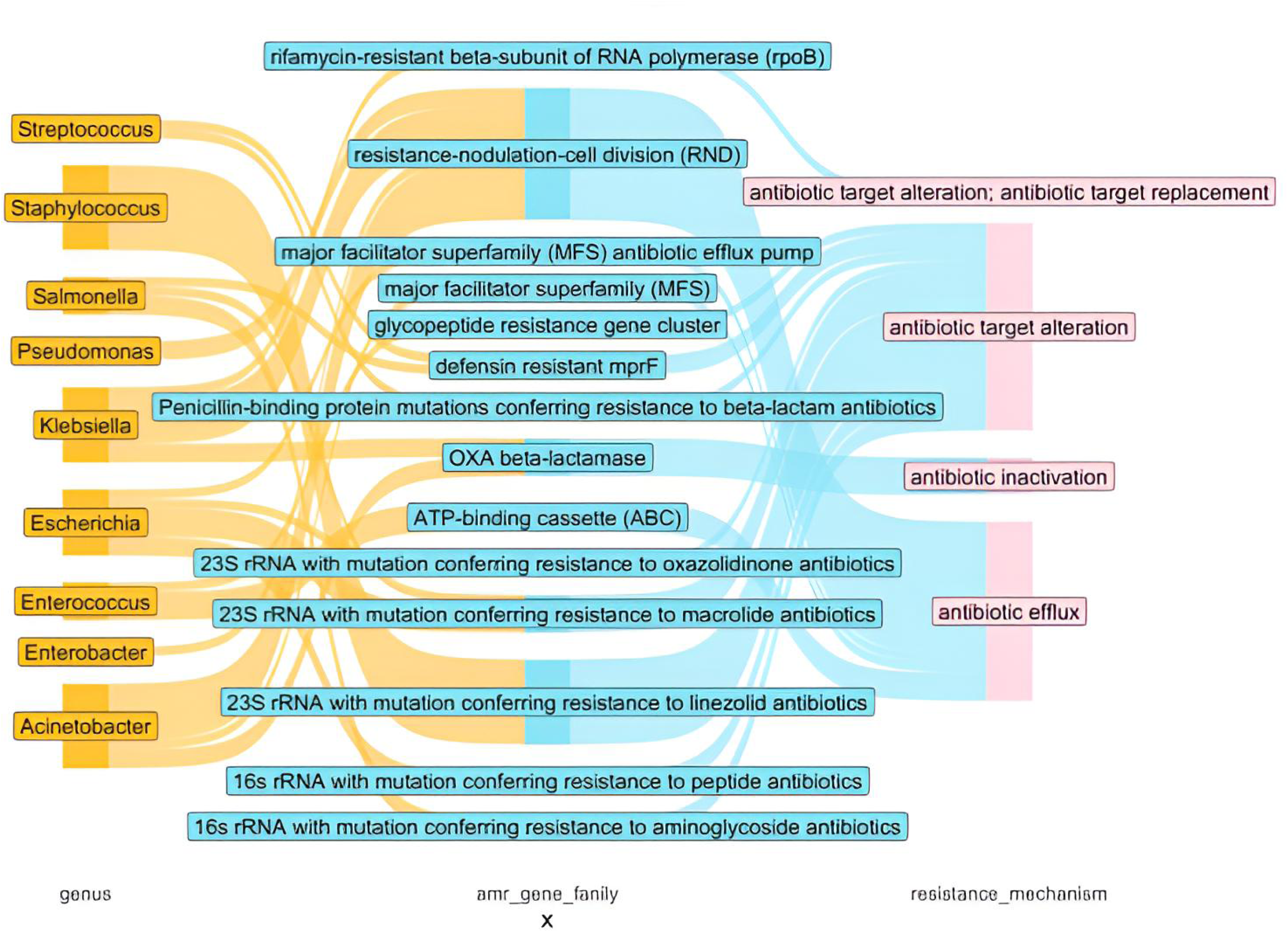
Sankey plot illustrating the relationships between sample IDs, pathogens detected by next-generation sequencing, antimicrobial resistance (AMR) gene families, and their associated resistance mechanisms across bloodstream infection samples.

AMR determinants were classified into three major resistance mechanisms: antibiotic inactivation, antibiotic efflux, and target alteration. Genes associated with antibiotic inactivation were predominantly β-lactamases, with OXA-type enzymes frequently detected in *Klebsiella* spp. and *Acinetobacter* spp. Additional β-lactamase families were identified across multiple Gram-negative genera.

Antibiotic efflux was the most frequently observed resistance mechanism. Resistance–nodulation–division (RND) efflux pumps were detected in *Acinetobacter*, *Klebsiella*, and *Pseudomonas*, while major facilitator superfamily (MFS) transporters were identified in *Acinetobacter* and *Escherichia*. ATP-binding cassette (ABC) transporters were observed among *Enterococcus* isolates. Additional efflux systems, including multidrug and toxic compound extrusion (MATE) and small multidrug resistance (SMR) families, were also detected across taxa.

Target alteration mechanisms were identified through ribosomal and protein-associated resistance determinants. Mutations in 23S rRNA associated with oxazolidinone resistance were detected in *Staphylococcus* spp., while macrolide resistance markers were identified in *Escherichia* and *Salmonella*. Mutations in 16S rRNA associated with aminoglycoside resistance were observed in *Salmonella*. Glycopeptide resistance gene clusters were detected in *Enterococcus*, and penicillin-binding protein–associated determinants were identified in *Streptococcus* spp.

### Plasmid Diversity among Bloodstream Infections

Metagenomic sequencing enabled identification of diverse plasmid replicons across bloodstream infection pathogens, including Inc-, Col-, and rep-family plasmids (Fig. 5). Distinct plasmid distribution patterns were observed across Gram-negative, Gram-positive, and fungal organisms. The complete plasmid dataset is provided in Supplementary File S4.

**Figure 5.**
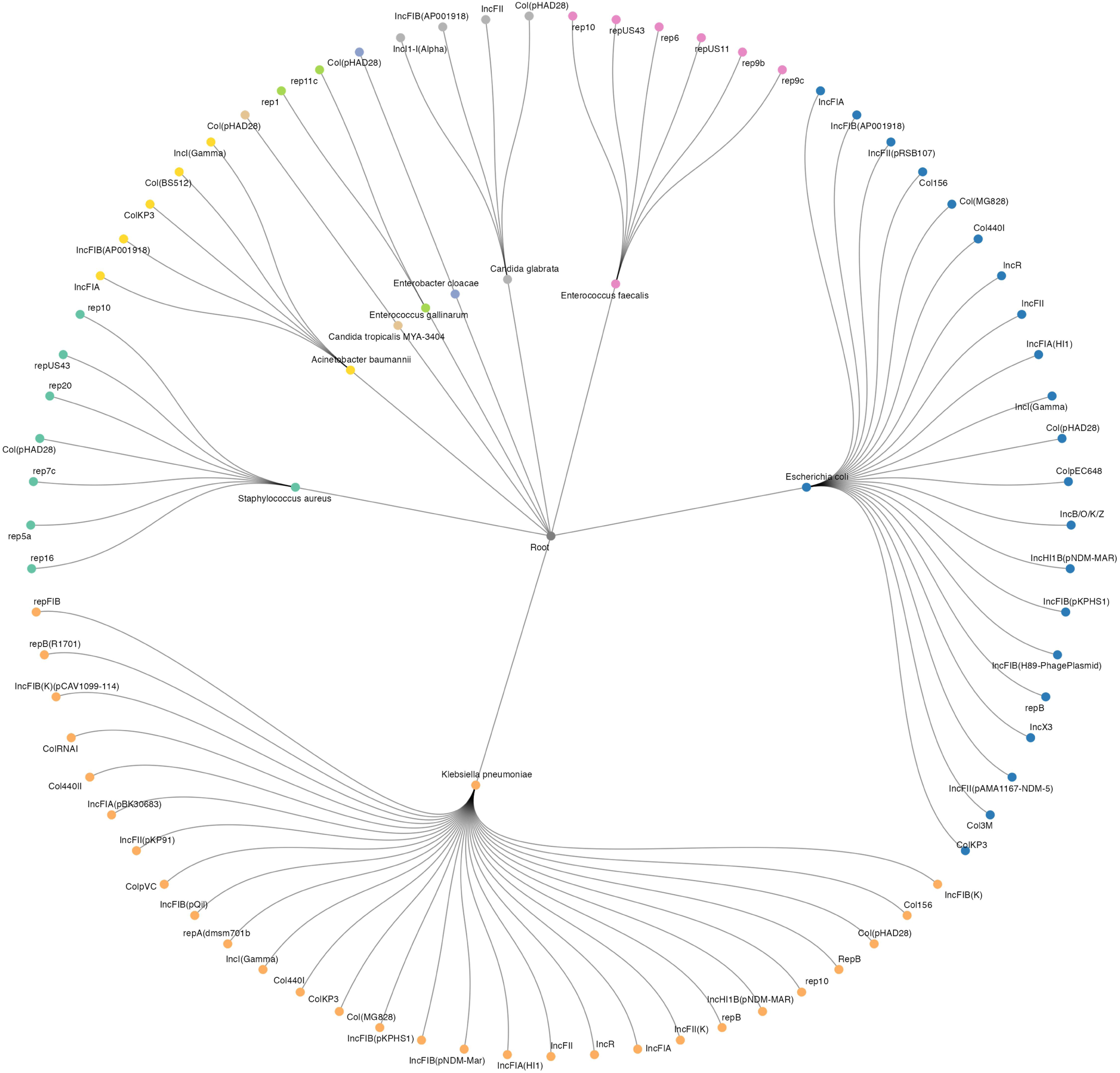
Detection and distribution of various plasmids across pathogens detected from clinical samples.

Among Gram-negative pathogens, *Klebsiella pneumoniae* and *Escherichia coli* exhibited the greatest plasmid diversity, with frequent detection of multiple Inc-family replicons, often co-occurring with Col-type plasmids. Rep-family plasmids, including *repA* and *repB* variants, were also identified in a subset of *K. pneumoniae* isolates. In contrast, *Acinetobacter baumannii* displayed a more limited plasmid repertoire, including IncFIA, IncFIB(AP001918), and IncI (Gamma) replicons, along with selected Col-type plasmids such as ColKP3 and Col(BS512). Rep-family plasmids were also detected in *A. baumannii*, comprising multiple rep subtypes. *Enterobacter cloacae* isolates predominantly carried Col(pHAD28).

Among Gram-positive pathogens, plasmid profiles were dominated by rep-family replicons. *Staphylococcus aureus* isolates harbored multiple rep-type plasmids, with additional Col-type plasmids detected in some samples. Similarly, *Enterococcus faecalis* and *Enterococcus gallinarum* carried multiple rep-family replicons.

Plasmid-associated sequences were also detected in fungal pathogens, including *Candida tropicalis* and *Candida glabrata*, where both Col-type and Inc-family plasmids were identified.

Overall, plasmid profiles varied across pathogen groups, with Inc- and Col-family plasmids more commonly observed among Enterobacterales and rep-family plasmids predominating among Gram-positive organisms.

## Discussion

Bloodstream infections (BSIs) remain time-critical clinical emergencies in which delays or inaccuracies in pathogen identification and antimicrobial resistance (AMR) profiling directly influence patient outcomes and antimicrobial stewardship (27). Although blood culture continues to serve as the diagnostic gold standard, its well-recognized limitations,including prolonged turnaround time, reduced sensitivity following empirical antibiotic exposure, and incomplete recovery of fastidious organisms,significantly constrain its effectiveness in routine clinical practice (28–30). In this study, we applied shotgun metagenomic next-generation sequencing (mNGS) directly to flagged positive blood culture bottles and evaluated its performance against phenotypic identification and antimicrobial susceptibility testing (AST). This paired framework enabled a robust assessment of concordance while also situating mNGS within a clinically relevant and implementable diagnostic pathway.

The taxonomic profiles observed in this cohort align with the expected epidemiology of BSIs in Indian tertiary care settings, with a predominance of Gram-negative pathogens. *Escherichia coli*, *Klebsiella pneumoniae*, and *Acinetobacter baumannii* were most frequently identified, followed by *Pseudomonas aeruginosa*, *Staphylococcus aureus*, and *Enterococcus* spp., consistent with the burden of ESKAPEE pathogens in healthcare-associated infections (31, 32). The high concordance between mNGS-based identification and routine blood culture supports the analytical robustness of sequencing from flagged blood culture material and reinforces its reliability within existing laboratory workflows.

Beyond concordance, mNGS consistently demonstrated improved species-level resolution compared with conventional phenotypic methods, particularly within genera such as *Acinetobacter*, *Enterococcus*, and *Streptococcus*. This added resolution is clinically meaningful, as closely related species may differ substantially in resistance profiles, virulence potential, and transmission dynamics (13, 33). The accurate identification of invasive fungal pathogens, including *Candida auris*, *Candida glabrata*, and *Candida tropicalis*, further highlights the value of unbiased sequencing in scenarios where delayed or inaccurate identification may directly impact therapeutic decision-making (13).

Although metagenomics is often emphasized for its ability to detect polymicrobial infections, sequencing from flagged blood culture bottles in this study predominantly reflected the dominant cultured organism. This observation reinforces the specificity conferred by the enrichment step inherent to blood culture workflows. At the same time, detection of low-abundance organisms, such as *Halomonas* spp., illustrates the sensitivity of mNGS to minor microbial signals. These findings require careful clinical interpretation to distinguish clinically relevant infection from background or transient DNA signals (34, 35).

A key translational advantage of this approach lies in its ability to recover pathogen genomic material despite prior antimicrobial exposure (36). In routine clinical settings, empirical antibiotics are frequently initiated before sampling, which can significantly reduce culture sensitivity (37). By leveraging the microbial enrichment achieved in flagged blood culture bottles, mNGS enables recovery of pathogen DNA at a clinically actionable stage without reliance on subculture (38). This is particularly relevant in critically ill and immunocompromised populations, where early empirical therapy is unavoidable but rapid therapeutic refinement remains essential (39, 40).

Integration of phenotypic AST with metagenomic resistance profiling demonstrated strong concordance across susceptibility categories (41). By restricting analysis to samples with a single cultured organism, this study enabled precise linkage of resistance determinants to their corresponding pathogens, allowing a controlled and clinically meaningful evaluation of performance. Extension of this framework to polymicrobial bloodstream infections will be an important next step to further define real-world applicability.

At a mechanistic level, resistance phenotypes in Gram-negative pathogens were supported by the detection of β-lactamases (including OXA-type and metallo-β-lactamases), aminoglycoside-modifying enzymes, quinolone resistance–associated mutations, and multidrug efflux systems (42). Comparable concordance was observed in Gram-positive organisms, including detection of methicillin resistance in *Staphylococcus aureus* and markers associated with macrolide and fluoroquinolone resistance (43, 44). These findings provide important biological context to phenotypic AST and strengthen confidence in resistance interpretation.

The detection of resistance-associated genes in phenotypically susceptible isolates likely reflects latent or inducible resistance mechanisms that may emerge under antimicrobial pressure (45, 46). Efflux-mediated resistance was the most prevalent mechanism observed, consistent with its central role in multidrug resistance across bacterial pathogens (47–49). Additional target modification mechanisms further contextualize resistance patterns (50, 51).

Together, these findings highlight the value of genomic data not only in confirming resistance but also in anticipating its potential evolution.

Plasmid profiling provided additional insight into the genetic basis of resistance dissemination. Inc- and Col-family plasmids predominated among Enterobacterales, consistent with their established role in horizontal gene transfer (52, 53), whereas rep-family plasmids were more common among Gram-positive organisms, suggesting lineage-associated resistance dynamics (54). While not immediately actionable at the individual patient level, these findings are highly relevant for infection control strategies and longitudinal AMR surveillance.

Collectively, these results support a clear and practical role for mNGS as an adjunct to conventional diagnostics. Beyond confirming pathogen identity, mNGS enhances diagnostic resolution, recovers clinically relevant information in settings where culture sensitivity is compromised,particularly following antibiotic exposure,and provides early, mechanistically grounded insight into antimicrobial resistance. In doing so, it strengthens the link between pathogen detection and resistance interpretation, a gap that remains a critical limitation of current diagnostic workflows.

Within the current diagnostic value chain, mNGS occupies an important intermediate position. In this study, its application downstream of blood culture, using flagged positive bottles, leverages microbial enrichment to generate high-confidence genomic data. More broadly, mNGS is increasingly being explored directly on primary clinical samples to enable earlier pathogen detection. These approaches are complementary: enrichment-based sequencing improves signal fidelity and interpretability, while direct-from-sample sequencing expands the scope of pathogen detection. However, widespread frontline implementation remains constrained by cost, infrastructure requirements, and the need for specialized analytical expertise, positioning mNGS currently as a high-resolution, reference-level and problem-solving tool.

Importantly, the genomic data generated in this study provide a rational framework for the development of targeted molecular diagnostics. By enabling the identification of robust species-specific and resistance-associated genetic markers, mNGS can guide the design of multiplex PCR assays that are more scalable, cost-effective, and suitable for routine deployment. This transition from discovery to deployment represents a critical translational pathway for integrating high-resolution genomic insights into accessible diagnostic platforms.

From a broader health systems perspective, these findings also have implications for AMR surveillance. Incorporating mNGS within diagnostic workflows offers the potential to enhance the sensitivity and resolution of surveillance systems, enabling earlier detection of resistance trends and more informed public health responses. While cost, infrastructure, and analytical complexity remain limiting factors (55–57), tiered implementation strategies,where mNGS supports reference-level analysis and informs downstream diagnostic development, may provide a pragmatic pathway toward scalable integration aligned with both clinical and public health priorities.

This study has several limitations, including its retrospective design, moderate sample size, and lack of detailed clinical outcome data. Prospective studies are needed to evaluate the impact of mNGS-guided diagnostics on antimicrobial decision-making, time to targeted therapy, and patient outcomes. Evaluation in polymicrobial infections and across diverse clinical settings will further strengthen the evidence base for implementation.

In summary, mNGS applied to flagged positive blood culture bottles reliably recapitulates culture-based pathogen identification and phenotypic antimicrobial susceptibility while providing additional genomic insight into resistance mechanisms and dissemination. These findings position mNGS as both a practical adjunct to conventional diagnostics and a translational bridge toward the development of targeted, scalable molecular assays. With continued validation, standardization, and integration into clinical and surveillance frameworks, mNGS holds strong potential to enhance diagnostic precision and support more responsive, data-driven approaches to bloodstream infection management and AMR control.

## Supporting information

Supplementary data for AST

Supplementary date Figure 2

Supplementary date Figure 3

Supplementary date Figure 4

## Acknowledgments

We thank Surabhi Srivastava (Tata Institute for Genetics and Society) for conceptual input and guidance on bioinformatics pipelines; Divya Tej Sowpati (Centre for Cellular and Molecular Biology) for assistance with bioinformatics workflows; and Awadhesh Pandit (National Centre for Biological Sciences) for support with sequencing and data backup. We acknowledge L. S. Shashidhara (National Centre for Biological Sciences) for facilitating connections with Bangalore Baptist Hospital and for his encouragement, and we thank the staff of Bangalore Baptist Hospital, Bengaluru, and Tata Institute for Genetics and Society for their support. This work was supported by Tata Trusts and Rockefeller Foundation (grant no. 2021 HTH018).

## Data Availability

Raw shotgun metagenomic sequencing reads generated from 55 flagged positive blood culture bottles in this study have been deposited in the NCBI Sequence Read Archive (SRA) under BioProject accession number PRJNA1459685, with individual BioSample accessions SAMN57562785–SAMN57562839. A complete mapping of sample identifiers to BioSample and SRA accessions is provided in Supplementary Table S5. Antimicrobial resistance gene profiles, plasmid replicon data, and per-sample taxonomic abundance data supporting the findings of this study are provided in Supplementary Files S1–S4.

