## Supplementary data for AST for "Deciphering antimicrobial resistance in bloodstream infections through clinical metagenomics"


**BLOOD CULTURE**

**ANTIBIOTIC PATTERN**

| Sample ID | Short ID | Sample ID BBH | Type of Sample | Nature of Sample | Sample Type | Type of Organism | Hospital Detected | Ampicillin (DISC/ | Gentamicin-Syn | Linezolid (DISC/M | Penicillin (DISC/M | Teicoplanin (DISC | Vancomycin (DISC/MIC) |
|---|---|---|---|---|---|---|---|---|---|---|---|---|---|
| BB30 | S61 | B-3109-24 | Blood | Blood Bottle | DNA | Human | Enterococcus sp | S | S | S | S | S | S |
| BB55 | S15 | B-5227/24 | Blood | Blood Bottle | DNA | Human | Enterococcus faecalis | S | R | S | S | _ | S |
| BB82 | S32 | B-6208/24 | Blood | Blood Bottle | DNA | Human | Enterococcus faecalis | S | S | S | S | S | S |
| BB157 | S45 | B-8339-24 | Blood | Blood Bottle | DNA | Human | Enterococcus sp | S | S | S | S | S | R |

| | | | | | | | | Amikacin (DISC/M | Ampicillin (DISC/ | Cefazolin (DISC/ | Cefipime (DISC/M | Cefoperazone/Su | Ceftriaxone (DIS | Cefuroxime (DIS | Ciprofloxacin (DI | Co-trimoxazole ( | Ertapenam (DISC | Gentamicin (DIS | Imipenem (DISC/ | Meropenem DIS | Piperacillin/Tazobactum (DISC/MIC) |
|---|---|---|---|---|---|---|---|---|---|---|---|---|---|---|---|---|---|---|---|---|---|
| BB43 | S9 | B-4245/24 | Blood | Blood Bottle | DNA | Human | Enterobacter sp | S | R | R | S | S | S | R | S | S | S | S | S | S | S |

| | | | | | | | | Ampicillin (DISC) | Cefotaxime (DIS | Ceftriaxone (DIS | Clindamycin (DIS | Co-trimoxazole ( | Erythromycin (DI | Levofloxacin (DIS | Linezolid (DISC/M | Penicillin (DISC/M | Vancomycin (DISC/MIC) |
|---|---|---|---|---|---|---|---|---|---|---|---|---|---|---|---|---|---|
| BB47 | S251 | B-4339/24 | Blood | Blood Bottle | DNA | Human | Streptococcus agalactiae | _ | _ | S | S | _ | S | S | S | S | S |
| BB48 | S252 | B-4516/24 | Blood | Blood Bottle | DNA | Human | Streptococcus pyogenes | S | S | _ | R | _ | R | I | S | S | S |
| BB162 | S48 | B-8558-24 | Blood | Blood Bottle | DNA | Human | Streptococcus pneumoniae | _ | S | S | S | R | R | S | S | S | S |

| | | | | | | | | Amikacin (DISC/M | Cefipime (DISC/M | Cefoperazone/Su | Cefotaxime (DIS | Ceftazidime (DIS | Ceftriaxone (DIS | Ciprofloxacin (DI | Colistin (DISC/MI | Co-trimoxazole ( | Gentamicin (DIS | Imipenem (DISC/ | Levofloxacin (DIS | Meropenem (DIS | Piperacillin/Tazobactum (DISC/MIC) |
|---|---|---|---|---|---|---|---|---|---|---|---|---|---|---|---|---|---|---|---|---|---|
| BB36 | S7 | B-3993-24 | Blood | Blood Bottle | DNA | Human | Acinetobacter | I | R | I | R | R | R | R | I | S | R | R | R | R | R |
| BB50 | S12 | B-4954-24 | Blood | Blood Bottle | DNA | Human | Acinetobacter | I | R | R | R | R | R | R | I | R | S | R | I | R | R |
| BB57 | S16 | B-5286/24 | Blood | Blood Bottle | DNA | Human | Acinetobacter | R | R | R | R | R | R | R | I | R | R | R | R | R | R |
| BB60 | S17 | B-5295/24 | Blood | Blood Bottle | DNA | Human | Acinetobacter | R | R | R | R | R | R | R | I | S | R | R | R | R | R |
| BB61 | S18 | B-5163/24 | Blood | Blood Bottle | DNA | Human | Acinetobacter baumannii (MDR) | R | R | R | _ | _ | R | R | I | R | R | R | _ | R | R |
| BB72 | S26 | B-5996/24 | Blood | Blood Bottle | DNA | Human | Acinetobacter | R | R | I | R | R | R | R | I | R | R | R | R | R | R |
| BB79 | S30 | B-6058/24 | Blood | Blood Bottle | DNA | Human | Acinetobacter | R | I | R | R | R | R | R | I | R | R | I | R | S | R |
| BB85 | S33 | B-6196/24 | Blood | Blood Bottle | DNA | Human | Acinetobacter baumannii complex | I | S | S | S | S | S | S | I | S | S | S | S | S | R |
| BB112 | S37 | B-7004-24 | Blood | Blood Bottle | DNA | Human | Acinetobacter | S | R | S | R | R | I | S | I | R | I | R | S | S | R |
| BB118 | S38 | B-7193-24 | Blood | Blood Bottle | DNA | Human | Acinetobacter | R | R | R | R | R | R | R | I | R | R | R | R | R | R |

| | | | | | | | | Amikacin (DISC/M | Ampicillin (DISC/ | Cefazolin (DISC/ | Cefipime (DISC/M | Cefoperazone/Su | Ceftriaxone (DIS | Cefuroxime (DIS | Ciprofloxacin (DI | Colistin (DISC/M | Co-trimoxazole ( | Ertapenam DISC | Gentamicin (DIS | Imipenem (DISC/ | Meropenem (DIS | Piperacillin /Tazo | Tigecycline (DISC/MIC) |
|---|---|---|---|---|---|---|---|---|---|---|---|---|---|---|---|---|---|---|---|---|---|---|---|
| BB31 | S248 | B-3072-24 | Blood | Blood Bottle | DNA | Human | E.coli | S | R | R | _ | S | R | R | R | _ | R | S | S | S | S | S | _ |
| BB54 | S14 | B-5257/24 | Blood | Blood Bottle | DNA | Human | E.coli | S | R | R | R | S | R | R | I | I | S | S | S | S | S | S | S |
| BB59 | S253 | B-5062/24 | Blood | Blood Bottle | DNA | Human | E.coli | S | R | R | R | S | R | R | R | I | S | S | S | S | S | S | S |
| BB64 | S20 | B-5482/24 | Blood | Blood Bottle | DNA | Human | E.coli | S | R | R | R | S | R | R | I | I | S | S | S | S | S | S | S |
| BB65 | S21 | B-5462/24 | Blood | Blood Bottle | DNA | Human | E.coli | S | R | R | SDD | R | R | R | R | I | R | I | R | S | S | R | S |
| BB76 | S29 | B-5770/24 | Blood | Blood Bottle | DNA | Human | E.coli | I | _ | R | R | S | R | R | R | I | R | S | R | S | S | S | S |
| BB115 | S255 | B-7334-24 | Blood | Blood Bottle | DNA | Human | E.coli | S | R | R | R | R | R | R | R | I | R | R | S | R | R | R | S |
| BB151 | S43 | B-8157-24 | Blood | Blood Bottle | DNA | Human | E.coli | S | R | R | R | S | R | R | R | I | R | S | S | S | S | S | S |

| | | | | | | | | Amoxicillin-clavul | Cefoxitin (DISC/M | Cefuroxime (Bas | Cephalexin (Bas | Chloramphenicol | Ciprofloxacin (DI | Clindamycin (DIS | Co-trimoxazole ( | Erythromycin (DI | Levofloxacin (DIS | Linezolid (DISC/M | Teicoplanin (DIS | Tetracycline (DIS | Vancomycin (DISC/MIC) |
|---|---|---|---|---|---|---|---|---|---|---|---|---|---|---|---|---|---|---|---|---|---|
| BB35 | S6 | B-3190-24 | Blood | Blood Bottle | DNA | Human | Staphalococcus aureus MSSA | S | S | S | S | S | R | S | S | S | R | S | S | S | S |
| BB39 | S249 | B-3894-24 | Blood | Blood Bottle | DNA | Human | Staphalococcus aureus MRSA | R | R | R | R | S | R | S | S | S | R | S | S | S | S |
| BB45 | S250 | B-4637/24 | Blood | Blood Bottle | DNA | Human | Staphalococcus aureus MSSA | S | S | S | S | S | R | S | R | S | R | S | S | S | S |
| BB46 | S10 | B4643/24 | Blood | Blood Bottle | DNA | Human | Staphalococcus aureus MSSA | S | S | S | S | S | S | S | S | S | S | S | S | S | S |
| BB62 | S19 | B-5554/24 | Blood | Blood Bottle | DNA | Human | Staphalococcus aureus MRSA | R | R | R | R | S | R | S | S | R | R | S | S | S | S |
| BB68 | S254 | B-5518/24 | Blood | Blood Bottle | DNA | Human | Staphalococcus aureus MRSA | R | R | R | R | S | R | R | R | R | R | S | S | S | S |
| BB70 | S24 | B-5511/24 | Blood | Blood Bottle | DNA | Human | Staphalococcus aureus MRSA | R | R | R | R | S | R | S | R | R | R | S | S | S | S |
| BB71 | S25 | B-5666/24 | Blood | Blood Bottle | DNA | Human | Staphalococcus aureus MSSA | S | S | S | S | S | R | R | S | S | R | S | S | S | S |
| BB75 | S28 | B-5842/24 | Blood | Blood Bottle | DNA | Human | Staphalococcus aureus MRSA | R | R | R | R | S | R | S | S | R | R | S | S | S | S |
| BB160 | S46 | B-8501-24 | Blood | Blood Bottle | DNA | Human | Staphalococcus aureus MSSA | S | S | S | S | S | R | S | S | S | R | S | S | S | S |

| | | | | | | | | Amikacin (DISC/M | Ampicillin (DISC/ | Cefazolin (DISC/ | Cefipime (DISC/M | Cefoperazone/Su | Ceftriaxone (DIS | Cefuroxime (DIS | Ciprofloxacin (DI | Colistin (DISC/M | Co-trimoxazole ( | Ertapenam (DISC | Gentamicin (DIS | Imipenem (DISC/ | Meropenem (DIS | Piperacillin/Tazot | Tigecycline (DISC/MIC) |
|---|---|---|---|---|---|---|---|---|---|---|---|---|---|---|---|---|---|---|---|---|---|---|---|
| BB32 | S3 | B-3149-24 | Blood | Blood Bottle | DNA | Human | Klebsiella sp | S | R | S | _ | _ | S | S | S | _ | S | _ | S | S | S | S | _ |
| BB33 | S4 | B-3176-24 | Blood | Blood Bottle | DNA | Human | Klebsiella sp | S | R | S | S | S | S | S | S | _ | S | S | S | S | S | S | _ |
| BB37 | S8 | B-3934-24 | Blood | Blood Bottle | DNA | Human | Klebsiella pneumoniae | S | R | R | S | S | S | R | I | _ | S | S | S | S | S | S | _ |
| BB66 | S22 | B-5438/24 | Blood | Blood Bottle | DNA | Human | Klebsiella pneumoniae | S | R | S | S | S | S | S | S | I | S | S | S | S | S | S | S |
| BB67 | S23 | B-5528/24 | Blood | Blood Bottle | DNA | Human | Klebsiella pneumoniae | R | R | R | R | R | R | R | R | I | S | R | R | R | R | R | S |
| BB74 | S27 | B-5991/24 | Blood | Blood Bottle | DNA | Human | Klebsiella sp | S | R | R | R | R | R | R | R | I | R | R | S | R | R | R | R |
| BB81 | S31 | B-6190/24 | Blood | Blood Bottle | DNA | Human | Klebsiella pneumoniae | R | R | R | R | R | R | R | R | I | R | R | R | R | R | R | S |
| BB108 | S35 | B-6969-24 | Blood | Blood Bottle | DNA | Human | Klebsiella sp | S | R | R | R | S | R | R | R | I | R | S | R | S | S | S | S |

| | | | | | | | | Amikacin (DISC/M | Cefipime (DISC/M | Cefoperazone/Su | Ceftazidime (DIS | Ciprofloxacin (DI | Colistin (DISC/M | Imipenem (DISC/ | Levofloxacin (DIS | Meropenem (DIS | Piperacillin/Tazobactum (DISC/MIC) |
|---|---|---|---|---|---|---|---|---|---|---|---|---|---|---|---|---|---|
| BB34 | S5 | B-3219-24 | Blood | Blood Bottle | DNA | Human | Pseudomonas aeuroginosa | S | S | S | S | S | I | S | S | S | S |
| BB51 | S13 | B-4701-24 | Blood | Blood Bottle | DNA | Human | Pseudomonas aeuroginosa | S | S | S | S | S | I | S | S | S | S |
| BB87 | S44 | B-6303-24 | Blood | Blood Bottle | DNA | Human | Pseudomonas aeuroginosa | S | S | S | S | S | I | S | S | S | S |
| BB110 | S36 | B-6803-24 | Blood | Blood Bottle | DNA | Human | Pseudomonas aeuroginosa | S | S | S | S | S | I | S | S | S | S |

| | | | | | | | | Ampicillin (DISC/ | Azithromycin (DI | Ceftriaxone (DIS | Chloramphenicol | Ciprofloxacin (DI | Co-trimoxazole (DISC/MIC) |
|---|---|---|---|---|---|---|---|---|---|---|---|---|---|
| BB49 | S11 | B-4880-24 | Blood | Blood Bottle | DNA | Human | Salmonella enterica serotype typhi | S | S | S | S | I | S |
| BB126 | S39 | B-6948-24 | Blood | Blood Bottle | DNA | Human | Salmonella enterica serotype typhi(Typhoid ) | S | S | S | S | I | S |
| BB156 | S44 | B-8576-24 | Blood | Blood Bottle | DNA | Human | Salmonella enterica serotype typhi | S | S | S | S | I | S |
| BB161 | S47 | B-8555-24 | Blood | Blood Bottle | DNA | Human | Salmonella enterica serotype typhi | S | S | S | S | I | S |

| | | | | | | | | Amphotericin B ( | Caspofungin (DIS | Fluconazole (DIS | Flucytosine (DIS | Micafungin (DISC | Voriconazole (DISC/MIC) |
|---|---|---|---|---|---|---|---|---|---|---|---|---|---|
| BB136 | S40 | B-7358-24 | Blood | Blood Bottle | DNA | Human | Candida species | R | S | R | R | S | R |
| BB141 | S41 | B-7895-24 | Blood | Blood Bottle | DNA | Human | Candida species | S | S | _ | S | S | S |
| BB150 | S42 | B-8138-24 | Blood | Blood Bottle | DNA | Human | Candida tropicalis | S | S | S | S | S | S |
