## Supplementary date Figure 2 for "Deciphering antimicrobial resistance in bloodstream infections through clinical metagenomics"

**Fig 2. Relative abundance of bacterial and fungal species identified in positive blood culture samples (n = 55) using mNGS.**

| genus | species | relative_abundance | sample_id |
|---|---|---|---|
| Acinetobacter | Acinetobacter haemolyticus | 44.64 | S33 |
| Acinetobacter | Acinetobacter baumannii | 16.17 | S12 |
| Acinetobacter | Acinetobacter guillouiae | 14.45 | S37 |
| Acinetobacter | Acinetobacter baumannii | 5.49 | S7 |
| Acinetobacter | Acinetobacter baumannii | 5 | S18 |
| Acinetobacter | Acinetobacter baumannii | 4.99 | S16 |
| Acinetobacter | Acinetobacter baumannii | 4.44 | S17 |
| Acinetobacter | Acinetobacter baumannii | 4.41 | S38 |
| Acinetobacter | Acinetobacter baumannii | 4.05 | S26 |
| Candida | Candida glabrata | 84.68 | S41 |
| Candida | Candida auris | 29.25 | S40 |

| | | | |
|---|---|---|---|
| Candida | Candida tropicalis MYA-3404 | 14.58 | S42 |
| Enterobacter | Enterobacter cloacae | 13.69 | S9 |
| Enterococcus | Enterococcus faecalis | 22.46 | S61 |
| Enterococcus | Enterococcus faecalis | 21.75 | S32 |
| Enterococcus | Enterococcus faecalis | 20.38 | S15 |
| Enterococcus | Enterococcus gallinarum | 6.05 | S45 |
| Escherichia | Escherichia coli | 13.11 | S14 |
| Escherichia | Escherichia coli | 12.14 | S248 |
| Escherichia | Escherichia coli | 7.48 | S20 |
| Escherichia | Escherichia coli | 4.29 | S255 |
| Escherichia | Escherichia coli | 3.74 | S253 |
| Escherichia | Escherichia coli | 3.72 | S43 |
| Escherichia | Escherichia coli | 3.5 | S21 |
| Escherichia | Escherichia coli | 3.23 | S29 |
| Halomonas | Halomonas hydrothermalis | 40.49 | S30 |
| Klebsiella | Klebsiella pneumoniae | 11.19 | S8 |
| Klebsiella | Klebsiella pneumoniae | 9.05 | S3 |
| Klebsiella | Klebsiella pneumoniae | 8.94 | S22 |
| Klebsiella | Klebsiella pneumoniae | 8.49 | S4 |
| Klebsiella | Klebsiella pneumoniae | 7.65 | S35 |
| Klebsiella | Klebsiella pneumoniae | 7.64 | S23 |
| Klebsiella | Klebsiella pneumoniae | 7.14 | S31 |
| Klebsiella | Klebsiella pneumoniae | 3.62 | S27 |
| Pseudomonas | Pseudomonas aeruginosa | 6.57 | S13 |
| Pseudomonas | Pseudomonas aeruginosa | 5.75 | S5 |
| Pseudomonas | Pseudomonas aeruginosa | 4.58 | S36 |
| Pseudomonas | Psuedomonas aueroginosa | 5.99 | S34 |
| Salmonella | Salmonella enterica | 38.75 | S44 |
| Salmonella | Salmonella enterica | 38.53 | S47 |
| Salmonella | Salmonella enterica | 38.03 | S11 |

| | | | |
|---|---|---|---|
| Salmonella | Salmonella enterica | 37.81 | S39 |
| Staphylococcus | Staphylococcus aureus | 19.48 | S6 |
| Staphylococcus | Staphylococcus aureus | 11.17 | S28 |
| Staphylococcus | Staphylococcus aureus | 9.78 | S25 |
| Staphylococcus | Staphylococcus aureus | 8.78 | S46 |
| Staphylococcus | Staphylococcus aureus | 8.17 | S250 |
| Staphylococcus | Staphylococcus aureus | 7.69 | S249 |
| Staphylococcus | Staphylococcus aureus | 7.65 | S254 |
| Staphylococcus | Staphylococcus aureus | 6.59 | S10 |
| Staphylococcus | Staphylococcus aureus | 5.67 | S24 |
| Staphylococcus | Staphylococcus aureus | 5.29 | S19 |
| Streptococcus | Streptococcus pyogenes | 65.03 | S252 |
| Streptococcus | Streptococcus pneumoniae | 38.15 | S48 |
| Streptococcus | Streptococcus agalactiae | 19.39 | S251 |
