## Supplementary date Figure 3 for "Deciphering antimicrobial resistance in bloodstream infections through clinical metagenomics"

**Figure 3A AND 3B. Integrated heatmap of pathogen identity, antimicrobial drug class, phenotypic susceptibility (AST), and antimicrobial resistance gene (ARG) burden across bloodstream infection isolates.**

| **sample_id** | **genus** | **amr_gene_fanily** | **resistance_mechanism** |
|---|---|---|---|
| S33 | Acinetobacter | OXA beta-lactamase | antibiotic inactivation |
| S12 | Acinetobacter | resistance-nodulation-cell division (RND) | antibiotic efflux |
| S37 | Acinetobacter | OXA beta-lactamase | antibiotic inactivation |
| S7 | Acinetobacter | resistance-nodulation-cell division (RND) | antibiotic efflux |
| S18 | Acinetobacter | resistance-nodulation-cell division (RND) | antibiotic efflux |
| S16 | Acinetobacter | resistance-nodulation-cell division (RND) | antibiotic efflux |
| S17 | Acinetobacter | major facilitator superfamily (MFS) | antibiotic efflux |
| S38 | Acinetobacter | resistance-nodulation-cell division (RND) | antibiotic efflux |
| S26 | Acinetobacter | resistance-nodulation-cell division (RND) | antibiotic efflux |
| S9 | Enterobacter | resistance-nodulation-cell division (RND) | antibiotic efflux |
| S61 | Enterococcus | ATP-binding cassette (ABC) | antibiotic efflux |
| S32 | Enterococcus | ATP-binding cassette (ABC) | antibiotic efflux |
| S15 | Enterococcus | ATP-binding cassette (ABC) | antibiotic efflux |
| S45 | Enterococcus | glycopeptide resistance gene cluster | antibiotic target alteration |
| S14 | Escherichia | 16s rRNA with mutation conferring resistance to peptide antibiotics | antibiotic target alteration |
| S20 | Escherichia | 23S rRNA with mutation conferring resistance to macrolide antibiotics | antibiotic target alteration |
| S255 | Escherichia | 23S rRNA with mutation conferring resistance to oxazolidinone antibiotics | antibiotic target alteration |
| S253 | Escherichia | major facilitator superfamily (MFS) antibiotic efflux pump | antibiotic efflux |
| S43 | Escherichia | 23S rRNA with mutation conferring resistance to macrolide antibiotics | antibiotic target alteration |
| S21 | Escherichia | 23S rRNA with mutation conferring resistance to macrolide antibiotics | antibiotic target alteration |
| S29 | Escherichia | 16s rRNA with mutation conferring resistance to peptide antibiotics | antibiotic target alteration |
| S8 | Klebsiella | rifamycin-resistant beta-subunit of RNA polymerase (rpoB) | antibiotic target alteration; antibiotic target replacement |
| S3 | Klebsiella | resistance-nodulation-cell division (RND) | antibiotic efflux |
| S22 | Klebsiella | resistance-nodulation-cell division (RND) | antibiotic efflux |
| S4 | Klebsiella | resistance-nodulation-cell division (RND) | antibiotic efflux |
| S35 | Klebsiella | resistance-nodulation-cell division (RND) | antibiotic efflux |
| S23 | Klebsiella | OXA beta-lactamase | antibiotic inactivation |
| S31 | Klebsiella | resistance-nodulation-cell division (RND) | antibiotic efflux |
| S27 | Klebsiella | OXA beta-lactamase | antibiotic inactivation |

| | | | |
|---|---|---|---|
| S5 | Pseudomonas | resistance-nodulation-cell division (RND) | antibiotic efflux |
| S34 | Pseudomonas | resistance-nodulation-cell division (RND) | antibiotic efflux |
| S44 | Salmonella | 23S rRNA with mutation conferring resistance to macrolide antibiotics | antibiotic target alteration |
| S47 | Salmonella | defensin resistant mprF | antibiotic target alteration |
| S11 | Salmonella | 16s rRNA with mutation conferring resistance to aminoglycoside antibiotics | antibiotic target alteration |
| S39 | Salmonella | 16s rRNA with mutation conferring resistance to aminoglycoside antibiotics | antibiotic target alteration |
| S6 | Staphylococcus | 23S rRNA with mutation conferring resistance to linezolid antibiotics | antibiotic target alteration |
| S28 | Staphylococcus | 23S rRNA with mutation conferring resistance to linezolid antibiotics | antibiotic target alteration |
| S25 | Staphylococcus | 23S rRNA with mutation conferring resistance to linezolid antibiotics | antibiotic target alteration |
| S46 | Staphylococcus | 23S rRNA with mutation conferring resistance to linezolid antibiotics | antibiotic target alteration |
| S250 | Staphylococcus | 23S rRNA with mutation conferring resistance to linezolid antibiotics | antibiotic target alteration |
| S249 | Staphylococcus | 23S rRNA with mutation conferring resistance to linezolid antibiotics | antibiotic target alteration |
| S10 | Staphylococcus | 23S rRNA with mutation conferring resistance to linezolid antibiotics | antibiotic target alteration |
| S24 | Staphylococcus | 23S rRNA with mutation conferring resistance to linezolid antibiotics | antibiotic target alteration |
| S19 | Staphylococcus | 23S rRNA with mutation conferring resistance to linezolid antibiotics | antibiotic target alteration |
| S252 | Streptococcus | Penicillin-binding protein mutations conferring resistance to beta-lactam antibiotics | antibiotic target alteration |
| S251 | Streptococcus | defensin resistant mprF | antibiotic target alteration |
