## Supplementary date Figure 4 for "Deciphering antimicrobial resistance in bloodstream infections through clinical metagenomics"

**Figure 4. Sankey plot illustrating the relationships between sample IDs, pathogens detected by next-generation sequencing, antimicrobial resistance (AMR) gene families, and their associated resistance mechanisms across bloodstream infection samples**

| Sample ID | sample_id | Hospital Detected | Antibiotic | Pattern | ARO | Category |
|---|---|---|---|---|---|---|
| BB112 | S37 | Acinetobacter sp | Amikacin | S | | |
| BB112 | S37 | Acinetobacter sp | Cefipime | R | OXA-668, NDM-1 | penicillin beta-lactam |
| BB112 | S37 | Acinetobacter sp | Cefotaxime | R | NDM-1 | cephalosporin |
| BB112 | S37 | Acinetobacter sp | Ceftazidime | R | NDM-1 | cephalosporin |
| BB112 | S37 | Acinetobacter sp | Ceftriaxone | I | NDM-1 | cephalosporin |
| BB112 | S37 | Acinetobacter sp | Ciprofloxacin | S | | |
| BB112 | S37 | Acinetobacter sp | Co-trimoxazole | R | | |
| BB112 | S37 | Acinetobacter sp | Colistin | _ | | |
| BB112 | S37 | Acinetobacter sp | Gentamicin | I | | |
| BB112 | S37 | Acinetobacter sp | Imipenem | R | OXA-668 | carbapenem |
| BB112 | S37 | Acinetobacter sp | Meropenem | S | NDM-1 | carbapenem |
| BB112 | S37 | Acinetobacter sp | Piperacillin/Tazobactum | R | OXA-668, NDM-1 | |
| BB112 | S37 | Acinetobacter sp | Tigecycline | _ | | glycylcycline |
| BB118 | S38 | Acinetobacter sp | Amikacin | R | ANT(3")-IIc | aminoglycoside antibiotic |
| BB118 | S38 | Acinetobacter sp | Cefipime | R | OXA-66, NDM-1, Acinetobacter baumannii AbuO, adeJ, adeK, adeI, adeN | penicillin beta-lactam |
| BB118 | S38 | Acinetobacter sp | Cefotaxime | R | NDM-1, Acinetobacter baumannii AbuO, adeJ, adeK, adeI, adeN | cephalosporin |
| BB118 | S38 | Acinetobacter sp | Ceftazidime | R | NDM-1, Acinetobacter baumannii AbuO, adeJ, adeK, adeI, adeN | cephalosporin |
| BB118 | S38 | Acinetobacter sp | Ceftriaxone | R | NDM-1, Acinetobacter baumannii AbuO, adeJ, adeK, adeI, adeN | cephalosporin |
| BB118 | S38 | Acinetobacter sp | Ciprofloxacin | R | adeF, Acinetobacter baumannii gyrA conferring resistance to fluoroquinolones, Acinetobacter baumannii parC conferring resistance to fluoroquinolones, [illegible] | fluoroquinolone antibiotic |
| BB118 | S38 | Acinetobacter sp | Co-trimoxazole | R | sul2 | sulfonamide antibiotic |
| BB118 | S38 | Acinetobacter sp | Colistin | I | | |
| BB118 | S38 | Acinetobacter sp | Gentamicin | R | Acinetobacter baumannii AbuO, ANT(3")-IIc | aminoglycoside antibiotic |
| BB118 | S38 | Acinetobacter sp | Imipenem | R | NDM-1 | carbapenem |
| BB118 | S38 | Acinetobacter sp | Meropenem | R | OXA-66, Acinetobacter baumannii AbuO, adeJ, adeK, adeI, adeN | carbapenem |
| BB118 | S38 | Acinetobacter sp | Piperacillin/Tazobactum | R | adeJ, adeK, NDM-1, Acinetobacter baumannii AbuO, adeI, OXA-66, adeN | |
| BB118 | S38 | Acinetobacter sp | Tigecycline | S | adeB, Acinetobacter baumannii AbuO, adeA, adeR | glycylcycline |
| BB36 | S7 | Acinetobacter sp | Amikacin | I | ANT(2")-Ia, Acinetobacter baumannii AbuO, APH(3')-VIa | aminoglycoside antibiotic |
| BB36 | S7 | Acinetobacter sp | Cefipime | R | OXA-23, OXA-91, Acinetobacter baumannii AbuO, adeK, adeI, adeN | penicillin beta-lactam |
| BB36 | S7 | Acinetobacter sp | Cefotaxime | R | Acinetobacter baumannii AbuO, adeJ, adeK, adeI, adeN | cephalosporin |
| BB36 | S7 | Acinetobacter sp | Ceftazidime | R | Acinetobacter baumannii AbuO, adeJ, adeK, adeI, adeN | cephalosporin |
| BB36 | S7 | Acinetobacter sp | Ceftriaxone | R | Acinetobacter baumannii AbuO, adeJ, adeK, adeI, adeN | cephalosporin |
| BB36 | S7 | Acinetobacter sp | Ciprofloxacin | R | Acinetobacter baumannii parC conferring resistance to fluoroquinolones, adeF, Acinetobacter baumannii gyrA conferring resistance to fluoroquinolones, [illegible] | fluoroquinolone antibiotic |
| BB36 | S7 | Acinetobacter sp | Co-trimoxazole | S | | |
| BB36 | S7 | Acinetobacter sp | Colistin | I | | |
| BB36 | S7 | Acinetobacter sp | Gentamicin | R | ANT(2")-Ia, Acinetobacter baumannii AbuO, ANT(3")-IIc, APH(3')-VIa | aminoglycoside antibiotic |
| BB36 | S7 | Acinetobacter sp | Imipenem | R | adeJ | carbapenem |
| BB36 | S7 | Acinetobacter sp | Meropenem | R | CARB-16, AbuO, adeI, adeK, OXA-23, OXA-91, adeN | penicillin beta-lactam |
| BB36 | S7 | Acinetobacter sp | Piperacillin/Tazobactum | R | adeJ, CARB-16, Acinetobacter baumannii AbuO, adeK, adeI, OXA-23, OXA-91, adeN | |
| BB36 | S7 | Acinetobacter sp | Tigecycline | S | Acinetobacter baumannii AbuO, adeR | glycylcycline |
| BB50 | S12 | Acinetobacter sp | Amikacin | I | APH(3")-Ib | aminoglycoside antibiotic |
| BB50 | S12 | Acinetobacter sp | Cefipime | R | NDM-1, adeJ, adeI | penicillin beta-lactam |
| BB50 | S12 | Acinetobacter sp | Cefotaxime | R | NDM-1, adeJ, adeI | cephalosporin |

| | | | | | | |
|---|---|---|---|---|---|---|
| BB50 | S12 | Acinetobacter sp | Ceftazidime | R | NDM-1, adeJ, adeI | cephalosporin |
| BB50 | S12 | Acinetobacter sp | Ceftriaxone | R | NDM-1, adeJ, adeI | cephalosporin |
| BB50 | S12 | Acinetobacter sp | Ciprofloxacin | R | Acinetobacter baumannii gyrA conferring resistance to fluoroquinolones, Acinetobacter baumannii parC conferring resistance to fluoroquinolones, adeJ, a | fluoroquinolone antibiotic |
| BB50 | S12 | Acinetobacter sp | Co-trimoxazole | R | | |
| BB50 | S12 | Acinetobacter sp | Colistin | I | | |
| BB50 | S12 | Acinetobacter sp | Gentamicin | S | APH(3")-Ib | aminoglycoside antibiotic |
| BB50 | S12 | Acinetobacter sp | Imipenem | R | adeJ | carbapenem |
| BB50 | S12 | Acinetobacter sp | Meropenem | R | NDM-1, adeI | carbapenem |
| BB50 | S12 | Acinetobacter sp | Piperacillin/Tazobactum | R | adeJ, adeI, NDM-1 | |
| BB50 | S12 | Acinetobacter sp | Tigecycline | _ | | glycylcycline |
| BB57 | S16 | Acinetobacter sp | Amikacin | R | APH(6)-Id, Acinetobacter baumannii AbuO, ANT(3")-IIc, APH(3")-Ib | aminoglycoside antibiotic |
| BB57 | S16 | Acinetobacter sp | Cefipime | R | OXA-66, NDM-1, adeJ, adeK, adeI, adeN | penicillin beta-lactam |
| BB57 | S16 | Acinetobacter sp | Cefotaxime | R | NDM-1, Acinetobacter baumannii AbuO, adeJ, adeK, adeI, adeN | cephalosporin |
| BB57 | S16 | Acinetobacter sp | Ceftazidime | R | NDM-1, Acinetobacter baumannii AbuO, adeJ, adeK, adeI, adeN | cephalosporin |
| BB57 | S16 | Acinetobacter sp | Ceftriaxone | R | NDM-1, Acinetobacter baumannii AbuO, adeJ, adeK, adeI, adeN | cephalosporin |
| BB57 | S16 | Acinetobacter sp | Ciprofloxacin | R | adeF, Acinetobacter baumannii gyrA conferring resistance to fluoroquinolones, Acinetobacter baumannii parC conferring resistance to fluoroquinolones, a | fluoroquinolone antibiotic |
| BB57 | S16 | Acinetobacter sp | Co-trimoxazole | R | | |
| BB57 | S16 | Acinetobacter sp | Colistin | I | | |
| BB57 | S16 | Acinetobacter sp | Gentamicin | R | APH(6)-Id, Acinetobacter baumannii AbuO, ANT(3")-IIc, APH(3")-Ib | aminoglycoside antibiotic |
| BB57 | S16 | Acinetobacter sp | Imipenem | R | OXA-66 | carbapenem |
| BB57 | S16 | Acinetobacter sp | Meropenem | R | adeJ, Acinetobacter baumannii AbuO, NDM-1, adeI, adeN, adeK | carbapenem |
| BB57 | S16 | Acinetobacter sp | Piperacillin/Tazobactum | R | adeJ, adeK, Acinetobacter baumannii AbuO, NDM-1, adeI, OXA-66, adeN | |
| BB57 | S16 | Acinetobacter sp | Tigecycline | S | adeB, Acinetobacter baumannii AbuO, adeA, adeC, adeR | glycylcycline |
| BB60 | S17 | Acinetobacter sp | Amikacin | R | APH(6)-Id, Acinetobacter baumannii AbuO, ANT(3")-IIc, APH(3')-Ia, APH(3")-Ib | aminoglycoside antibiotic |
| BB60 | S17 | Acinetobacter sp | Cefipime | R | OXA-23, OXA-66, Acinetobacter baumannii AbuO, adeJ, adeK, adeN | penicillin beta-lactam |
| BB60 | S17 | Acinetobacter sp | Cefotaxime | R | ADC-25, Acinetobacter baumannii AbuO, adeJ, adeK, adeI, adeN | cephalosporin |
| BB60 | S17 | Acinetobacter sp | Ceftazidime | R | ADC-25, Acinetobacter baumannii AbuO, adeJ, adeK, adeI, adeN | cephalosporin |
| BB60 | S17 | Acinetobacter sp | Ceftriaxone | R | NDM-1, Acinetobacter baumannii AbuO, adeJ, adeK, adeI, adeN | cephalosporin |
| BB60 | S17 | Acinetobacter sp | Ciprofloxacin | R | adeF, Acinetobacter baumannii parC conferring resistance to fluoroquinolones, Acinetobacter baumannii gyrA conferring resistance to fluoroquinolones, a | fluoroquinolone antibiotic |
| BB60 | S17 | Acinetobacter sp | Co-trimoxazole | S | | |
| BB60 | S17 | Acinetobacter sp | Colistin | I | | |
| BB60 | S17 | Acinetobacter sp | Gentamicin | R | APH(6)-Id, Acinetobacter baumannii AbuO, ANT(3")-IIc, APH(3')-Ia, APH(3")-Ib | aminoglycoside antibiotic |
| BB60 | S17 | Acinetobacter sp | Imipenem | R | OXA-23 | carbapenem |
| BB60 | S17 | Acinetobacter sp | Meropenem | R | carO, OXA-66, Acinetobacter baumannii AbuO, adeJ, adeK, adeI, adeN | carbapenem |
| BB60 | S17 | Acinetobacter sp | Piperacillin/Tazobactum | R | adeJ, OXA-23, adeK, Acinetobacter baumannii AbuO, adeI, OXA-66, adeN | |
| BB60 | S17 | Acinetobacter sp | Tigecycline | S | Acinetobacter baumannii AbuO | glycylcycline |
| BB61 | S18 | Acinetobacter sp | Amikacin | R | | |
| BB61 | S18 | Acinetobacter sp | Cefipime | R | NDM-1, adeJ, adeK, adeI | penicillin beta-lactam |
| BB61 | S18 | Acinetobacter sp | Cefotaxime | - | NDM-1, adeJ, adeK, adeI | cephalosporin |
| BB61 | S18 | Acinetobacter sp | Ceftazidime | _ | NDM-1, adeJ, adeK, adeI | cephalosporin |
| BB61 | S18 | Acinetobacter sp | Ceftriaxone | R | NDM-1, adeJ, adeK, adeI | cephalosporin |
| BB61 | S18 | Acinetobacter sp | Ciprofloxacin | R | adeF, Acinetobacter baumannii gyrA conferring resistance to fluoroquinolones, Acinetobacter baumannii parC conferring resistance to fluoroquinolones, a | fluoroquinolone antibiotic |
| BB61 | S18 | Acinetobacter sp | Co-trimoxazole | R | sul2 | sulfonamide antibiotic |
| BB61 | S18 | Acinetobacter sp | Colistin | I | | |
| BB61 | S18 | Acinetobacter sp | Gentamicin | R | | |
| BB61 | S18 | Acinetobacter sp | Imipenem | R | NDM-1 | carbapenem |
| BB61 | S18 | Acinetobacter sp | Meropenem | R | adeJ, adeK, adeI | carbapenem |
| BB61 | S18 | Acinetobacter sp | Piperacillin/Tazobactum | R | adeJ | |
| BB61 | S18 | Acinetobacter sp | Tigecycline | _ | adeB, adeA, adeC | glycylcycline |
| BB72 | S26 | Acinetobacter sp | Amikacin | R | APH(6)-Id, Acinetobacter baumannii AbuO, ANT(3")-IIc, APH(3')-VIa, APH(3")-Ib | aminoglycoside antibiotic |
| BB72 | S26 | Acinetobacter sp | Cefipime | R | OXA-23, OXA-66, Acinetobacter baumannii AbuO, adeJ, adeK, adeI, adeN | penicillin beta-lactam |
| BB72 | S26 | Acinetobacter sp | Cefotaxime | R | Acinetobacter baumannii AbuO, adeJ, adeK, adeI, adeN | cephalosporin |
| BB72 | S26 | Acinetobacter sp | Ceftazidime | R | Acinetobacter baumannii AbuO, adeJ, adeK, adeI, adeN | cephalosporin |
| BB72 | S26 | Acinetobacter sp | Ceftriaxone | R | Acinetobacter baumannii AbuO, adeJ, adeK, adeI, adeN | cephalosporin |
| BB72 | S26 | Acinetobacter sp | Ciprofloxacin | R | Acinetobacter baumannii parC conferring resistance to fluoroquinolones, Acinetobacter baumannii gyrA conferring resistance to fluoroquinolones, adeG, | fluoroquinolone antibiotic |
| BB72 | S26 | Acinetobacter sp | Co-trimoxazole | R | | |
| BB72 | S26 | Acinetobacter sp | Colistin | I | | |
| BB72 | S26 | Acinetobacter sp | Gentamicin | R | APH(6)-Id, Acinetobacter baumannii AbuO, ANT(3")-IIc, APH(3')-VIa, APH(3")-Ib | aminoglycoside antibiotic |
| BB72 | S26 | Acinetobacter sp | Imipenem | R | OXA-23 | carbapenem |
| BB72 | S26 | Acinetobacter sp | Meropenem | R | OXA-66, Acinetobacter baumannii AbuO, adeJ, adeK, adeI, adeN | carbapenem |

| | | | | | | |
|---|---|---|---|---|---|---|
| BB72 | S26 | Acinetobacter sp | Piperacillin/Tazobactum | R | OXA-23, Acinetobacter baumannii AbuO, adeI, OXA-66, adeN | |
| BB72 | S26 | Acinetobacter sp | Tigecycline | _ | adeB, Acinetobacter baumannii AbuO, adeA, adeC, adeR | glycylcycline |
| BB79 | S30 | Acinetobacter sp | Amikacin | R | ANT(3")-IIa | aminoglycoside antibiotic |
| BB79 | S30 | Acinetobacter sp | Cefipime | I | OXA-10, VIM-2, NDM-1 | penicillin beta-lactam |
| BB79 | S30 | Acinetobacter sp | Cefotaxime | R | OXA-10, VIM-2, NDM-1 | cephalosporin |
| BB79 | S30 | Acinetobacter sp | Ceftazidime | R | OXA-10, VIM-2, NDM-1 | cephalosporin |
| BB79 | S30 | Acinetobacter sp | Ceftriaxone | R | OXA-10, VIM-2, NDM-1 | cephalosporin |
| BB79 | S30 | Acinetobacter sp | Ciprofloxacin | R | | |
| BB79 | S30 | Acinetobacter sp | Co-trimoxazole | R | | |
| BB79 | S30 | Acinetobacter sp | Colistin | I | | |
| BB79 | S30 | Acinetobacter sp | Gentamicin | R | ANT(3")-IIa | aminoglycoside antibiotic |
| BB79 | S30 | Acinetobacter sp | Imipenem | I | VIM-2 | carbapenem |
| BB79 | S30 | Acinetobacter sp | Meropenem | S | PME-1, NDM-1 | carbapenem |
| BB79 | S30 | Acinetobacter sp | Piperacillin/Tazobactum | R | VIM-2, NDM-1, OXA-10 | |
| BB79 | S30 | Acinetobacter sp | Tigecycline | S | | glycylcycline |
| BB85 | S33 | Acinetobacter sp | Amikacin | I | AAC(6')-Ig | aminoglycoside antibiotic |
| BB85 | S33 | Acinetobacter sp | Cefipime | S | OXA-214 | penicillin beta-lactam |
| BB85 | S33 | Acinetobacter sp | Cefotaxime | S | | |
| BB85 | S33 | Acinetobacter sp | Ceftazidime | S | | |
| BB85 | S33 | Acinetobacter sp | Ceftriaxone | S | | |
| BB85 | S33 | Acinetobacter sp | Ciprofloxacin | S | adeF | fluoroquinolone antibiotic |
| BB85 | S33 | Acinetobacter sp | Co-trimoxazole | S | | |
| BB85 | S33 | Acinetobacter sp | Colistin | _ | | |
| BB85 | S33 | Acinetobacter sp | Gentamicin | S | | |
| BB85 | S33 | Acinetobacter sp | Imipenem | S | OXA-214 | carbapenem |
| BB85 | S33 | Acinetobacter sp | Meropenem | S | | |
| BB85 | S33 | Acinetobacter sp | Piperacillin/Tazobactum | R | OXA-214 | |
| BB85 | S33 | Acinetobacter sp | Tigecycline | _ | | glycylcycline |
