## Supplementary data Figure 5 for "Deciphering antimicrobial resistance in bloodstream infections through clinical metagenomics"

**Figure 5. Detection and distribution of various plasmids across pathogens detected from clinical samples**

| sample_id | pathogen | plasmid | category_plasmid |
|---|---|---|---|
| S35 | Klebsiella pneumoniae | Col440I | col |
| S35 | Klebsiella pneumoniae | Col(pHAD28) | col |
| S35 | Klebsiella pneumoniae | IncFIB(K) | inc |
| S255 | Escherichia coli | Col(MG828) | col |
| S255 | Escherichia coli | Col(pHAD28) | col |
| S255 | Escherichia coli | ColpEC648 | col |
| S255 | Escherichia coli | IncFIA | inc |
| S255 | Escherichia coli | IncR | inc |
| S255 | Escherichia coli | IncFII | inc |
| S255 | Escherichia coli | IncFIA(HI1) | inc |
| S255 | Escherichia coli | IncI(Gamma) | inc |
| S43 | Escherichia coli | Col156 | col |
| S43 | Escherichia coli | IncFIA | inc |
| S43 | Escherichia coli | IncFIB(AP00191[illegible] | inc |
| S45 | Enterococcus gallinarum | rep1 | rep |
| S45 | Enterococcus gallinarum | rep11c | rep |
| S45 | Enterococcus gallinarum | rep11c | rep |
| S61 | Enterococcus faecalis | rep10 | rep |
| S61 | Enterococcus faecalis | repUS43 | rep |
| S3 | Klebsiella pneumoniae | Col156 | col |
| S3 | Klebsiella pneumoniae | Col(pHAD28) | col |
| S3 | Klebsiella pneumoniae | IncFIB(K) | inc |
| S3 | Klebsiella pneumoniae | repB | rep |
| S3 | Klebsiella pneumoniae | rep10 | rep |
| S4 | Klebsiella pneumoniae | IncHI1B(pNDM-[illegible] | inc |
| S4 | Klebsiella pneumoniae | repB | rep |
| S6 | Staphylococcus aureus | rep16 | rep |
| S6 | Staphylococcus aureus | rep5a | rep |
| S8 | Klebsiella pneumoniae | ColRNAI | col |
| S8 | Klebsiella pneumoniae | Col440I | col |
| S8 | Klebsiella pneumoniae | IncFIA(HI1) | inc |
| S8 | Klebsiella pneumoniae | IncFIB(K) | inc |
| S8 | Klebsiella pneumoniae | IncFIB(K)(pCAV1[illegible] | inc |

| | | | |
|---|---|---|---|
| S8 | Klebsiella pneumoniae | repB(R1701) | rep |
| S8 | Klebsiella pneumoniae | repFIB | rep |
| S8 | Klebsiella pneumoniae | RepB | rep |
| S249 | Staphylococcus aureus | rep7c | rep |
| S9 | Enterobacter cloacae | Col(pHAD28) | col |
| S250 | Staphylococcus aureus | Col(pHAD28) | col |
| S250 | Staphylococcus aureus | rep20 | rep |
| S251 | Staphylococcus aureus | repUS43 | rep |
| S14 | Escherichia coli | IncB/O/K/Z | inc |
| S14 | Escherichia coli | IncFIB(AP00191ε | inc |
| S14 | Escherichia coli | IncFII | inc |
| S15 | Enterococcus faecalis | repUS11 | rep |
| S15 | Enterococcus faecalis | rep9b | rep |
| S15 | Enterococcus faecalis | rep9c | rep |
| S253 | Escherichia coli | Col156 | col |
| S253 | Escherichia coli | Col(MG828) | col |
| S253 | Escherichia coli | IncFIA | inc |
| S253 | Escherichia coli | IncFIB(AP00191ε | inc |
| S253 | Escherichia coli | IncFII(pRSB107) | inc |
| S19 | Staphylococcus aureus | rep16 | rep |
| S19 | Staphylococcus aureus | rep5a | rep |
| S19 | Staphylococcus aureus | rep7c | rep |
| S20 | Escherichia coli | IncFII | inc |
| S20 | Escherichia coli | IncHI1B(pNDM-M | inc |
| S20 | Escherichia coli | IncFIB(pKPHS1) | inc |
| S20 | Escherichia coli | IncFIB(H89-Phag | inc |
| S20 | Escherichia coli | repB | rep |
| S21 | Escherichia coli | Col3M | col |
| S21 | Escherichia coli | ColKP3 | col |
| S21 | Escherichia coli | IncFIA | inc |
| S21 | Escherichia coli | IncFIB(AP00191ε | inc |
| S21 | Escherichia coli | IncX3 | inc |
| S21 | Escherichia coli | IncFII(pAMA1167 | inc |
| S21 | Escherichia coli | IncI(Gamma) | inc |
| S22 | Klebsiella pneumoniae | Col(pHAD28) | col |
| S22 | Klebsiella pneumoniae | IncFII(K) | inc |
| S22 | Klebsiella pneumoniae | IncFIB(K) | inc |
| S23 | Klebsiella pneumoniae | Col(MG828) | col |
| S23 | Klebsiella pneumoniae | ColKP3 | col |
| S23 | Klebsiella pneumoniae | Col440I | col |
| S23 | Klebsiella pneumoniae | Col(pHAD28) | col |
| S23 | Klebsiella pneumoniae | IncFIA | inc |
| S23 | Klebsiella pneumoniae | IncR | inc |
| S23 | Klebsiella pneumoniae | IncFII | inc |
| S23 | Klebsiella pneumoniae | IncHI1B(pNDM-M | inc |
| S23 | Klebsiella pneumoniae | IncFIA(HI1) | inc |
| S23 | Klebsiella pneumoniae | IncFIB(pNDM-Ma | inc |
| S23 | Klebsiella pneumoniae | IncFIB(pKPHS1) | inc |
| S23 | Klebsiella pneumoniae | IncI(Gamma) | inc |
| S23 | Klebsiella pneumoniae | repA(dmsm701b | rep |

| | | | |
|---|---|---|---|
| S24 | Staphylococcus aureus | rep16 | rep |
| S24 | Staphylococcus aureus | rep5a | rep |
| S25 | Staphylococcus aureus | rep16 | rep |
| S25 | Staphylococcus aureus | rep5a | rep |
| S26 | Acinetobacter baumannii | ColKP3 | col |
| S26 | Acinetobacter baumannii | Col(BS512) | col |
| S26 | Acinetobacter baumannii | IncFIA | inc |
| S26 | Acinetobacter baumannii | IncFIB(AP00191[illegible] | inc |
| S26 | Acinetobacter baumannii | IncI(Gamma) | inc |
| S27 | Klebsiella pneumoniae | ColpVC | col |
| S27 | Klebsiella pneumoniae | ColKP3 | col |
| S27 | Klebsiella pneumoniae | Col440I | col |
| S27 | Klebsiella pneumoniae | Col(pHAD28) | col |
| S27 | Klebsiella pneumoniae | IncFII | inc |
| S27 | Klebsiella pneumoniae | IncFIB(pQil) | inc |
| S28 | Staphylococcus aureus | rep16 | rep |
| S28 | Staphylococcus aureus | rep5a | rep |
| S29 | Escherichia coli | Col156 | col |
| S29 | Escherichia coli | Col440I | col |
| S29 | Escherichia coli | IncFIA | inc |
| S29 | Escherichia coli | IncFIB(AP00191[illegible] | inc |
| S31 | Klebsiella pneumoniae | Col440II | col |
| S31 | Klebsiella pneumoniae | Col(pHAD28) | col |
| S31 | Klebsiella pneumoniae | IncFII(K) | inc |
| S31 | Klebsiella pneumoniae | IncFIB(K) | inc |
| S31 | Klebsiella pneumoniae | IncFIB(pKPHS1) | inc |
| S31 | Klebsiella pneumoniae | IncFII(pKP91) | inc |
| S31 | Klebsiella pneumoniae | IncFIA(pBK3068[illegible] | inc |
| S32 | Enterococcus faecalis | rep6 | rep |
| S32 | Enterococcus faecalis | repUS43 | rep |
