## Supplementary data Table 1 for "Deciphering antimicrobial resistance in bloodstream infections through clinical metagenomics"

**Table 1: Genotypic and phenotypic detection of pathogens from flagged blood bottle samples and corresponding isolates**

| Sample_id | Genotypic detection (mNGS detection) | Phenotypic detection (Hospital detection) |
|---|---|---|
| S38 | Acinetobacter baumannii | Acinetobacter sp |
| S7 | Acinetobacter baumannii | Acinetobacter sp |
| S12 | Acinetobacter baumannii | Acinetobacter sp |
| S16 | Acinetobacter baumannii | Acinetobacter sp |
| S17 | Acinetobacter baumannii | Acinetobacter sp |
| S18 | Acinetobacter baumannii | Acinetobacter sp |
| S26 | Acinetobacter baumannii | Acinetobacter sp |
| S37 | Acinetobacter guillouiae | Acinetobacter sp |
| S33 | Acinetobacter haemolyticus | Acinetobacter sp |
| S40 | Candida auris | Candida sp |
| S41 | Candida glabrata | Candida sp |
| S42 | Candida tropicalis MYA-3404 | Candida tropicalis |
| S9 | Enterobacter cloacae | Enterobacter sp |
| S61 | Enterococcus faecalis | Enterococcus sp |
| S15 | Enterococcus faecalis | Enterococcus facalis |
| S32 | Enterococcus faecalis | Enterococcus facalis |
| S45 | Enterococcus gallinarum | Enterococcus sp |
| S255 | Escherichia coli | Escherichia coli |
| S43 | Escherichia coli | Escherichia coli |
| S248 | Escherichia coli | Escherichia coli |
| S14 | Escherichia coli | Escherichia coli |
| S253 | Escherichia coli | Escherichia coli |
| S20 | Escherichia coli | Escherichia coli |
| S21 | Escherichia coli | Escherichia coli |
| S29 | Escherichia coli | Escherichia coli |
| S30 | Halomonas hydrothermalis | Acinetobacter sp |
| S35 | Klebsiella pneumoniae | Klebsiella sp |
| S3 | Klebsiella pneumoniae | Klebsiella sp |
| S4 | Klebsiella pneumoniae | Klebsiella sp |
| S8 | Klebsiella pneumoniae | Klebsiella pneumonia |
| S22 | Klebsiella pneumoniae | Klebsiella pneumonia |
| S23 | Klebsiella pneumoniae | Klebsiella pneumonia |
| S27 | Klebsiella pneumoniae | Klebsiella sp |
| S31 | Klebsiella pneumoniae | Klebsiella pneumonia |
| S36 | Pseudomonas aeruginosa | Psuedomonas aueroginosa |
| S5 | Pseudomonas aeruginosa | Psuedomonas aueroginosa |

| | | |
|---|---|---|
| S13 | Pseudomonas aeruginosa | Psuedomonas aueroginosa |
| S34 | Pseudomonas aeruginosa | Psuedomonas aueroginosa |
| S39 | Salmonella enterica subsp. enterica serovar Typhi | Salmonella enterica serotype typhi(Typhoid ) |
| S44 | Salmonella enterica subsp. enterica serovar Typhi | Salmonella enterica serotype typhi |
| S47 | Salmonella enterica subsp. enterica serovar Typhi | Salmonella enterica serotype typhi |
| S11 | Salmonella enterica subsp. enterica serovar Typhi | Salmonella enterica serotype typhi |
| S46 | Staphylococcus aureus | Staphalococcus aureus MSSA |
| S6 | Staphylococcus aureus | Staphalococcus aureus MSSA |
| S10 | Staphylococcus aureus | Staphalococcus aureus MSSA |
| S19 | Staphylococcus aureus | Staphalococcus aureus MRSA |
| S249 | Staphylococcus aureus subsp. aureus | Staphalococcus aureus MRSA |
| S250 | Staphylococcus aureus subsp. aureus | Staphalococcus aureus MSSA |
| S251 | Staphylococcus aureus subsp. aureus | Streptococcus agalactiae |
| S254 | Staphylococcus aureus subsp. aureus | Staphalococcus aureus MRSA |
| S24 | Staphylococcus aureus subsp. aureus | Staphalococcus aureus MRSA |
| S25 | Staphylococcus aureus subsp. aureus | Staphalococcus aureus MSSA |
| S28 | Staphylococcus aureus subsp. aureus | Staphalococcus aureus MRSA |
| S48 | Streptococcus pneumoniae | Streptococcus peumonia |
| S252 | Streptococcus pyogenes | Streptococcus phyogenes |
