## Supplementary data Table 2 for "Deciphering antimicrobial resistance in bloodstream infections through clinical metagenomics"

| Accession | Sample Name | SPUID | Organism | Tax ID | Isolate |
|---|---|---|---|---|---|
| SAMN57562785 | S10 | S10 | Staphylococcus aureus | 1280 | Acinetobacter sp |
| SAMN57562786 | S11 | S11 | Salmonella enterica subsp. enterica serovar Typhi | 90370 | Acinetobacter sp |
| SAMN57562787 | S12 | S12 | Acinetobacter baumannii | 470 | Acinetobacter sp |
| SAMN57562788 | S13 | S13 | Pseudomonas aeruginosa | 287 | Acinetobacter sp |
| SAMN57562789 | S14 | S14 | Escherichia coli | 562 | Acinetobacter sp |
| SAMN57562790 | S15 | S15 | Enterococcus faecalis | 1351 | Acinetobacter sp |
| SAMN57562791 | S16 | S16 | Acinetobacter baumannii | 470 | Acinetobacter sp |
| SAMN57562792 | S17 | S17 | Acinetobacter baumannii | 470 | Acinetobacter sp |
| SAMN57562793 | S18 | S18 | Acinetobacter baumannii | 470 | Acinetobacter sp |
| SAMN57562794 | S19 | S19 | Staphylococcus aureus | 1280 | Candida sp |
| SAMN57562795 | S20 | S20 | Escherichia coli | 562 | Candida sp |
| SAMN57562796 | S21 | S21 | Escherichia coli | 562 | Candida tropicalis |
| SAMN57562797 | S22 | S22 | Klebsiella pneumoniae | 573 | Enterobacter sp |
| SAMN57562798 | S23 | S23 | Klebsiella pneumoniae | 573 | Enterococcus sp |
| SAMN57562799 | S24 | S24 | Staphylococcus aureus | 1280 | Enterococcus faecalis |
| SAMN57562800 | S248 | S248 | Escherichia coli | 562 | Enterococcus faecalis |
| SAMN57562801 | S249 | S249 | Staphylococcus aureus | 1280 | Enterococcus sp |
| SAMN57562802 | S25 | S25 | Staphylococcus aureus | 1280 | Escherichia coli |
| SAMN57562803 | S250 | S250 | Staphylococcus aureus | 1280 | Escherichia coli |
| SAMN57562804 | S251 | S251 | Staphylococcus aureus | 1280 | Escherichia coli |
| SAMN57562805 | S252 | S252 | Streptococcus pyogenes | 1314 | Escherichia coli |
| SAMN57562806 | S253 | S253 | Escherichia coli | 562 | not provided |
| SAMN57562807 | S254 | S254 | Staphylococcus aureus | 1280 | Escherichia coli |
| SAMN57562808 | S255 | S255 | Escherichia coli | 562 | not provided |
| SAMN57562809 | S26 | S26 | Acinetobacter baumannii | 470 | Escherichia coli |
| SAMN57562810 | S27 | S27 | Klebsiella pneumoniae | 573 | Acinetobacter sp |
| SAMN57562811 | S28 | S28 | Staphylococcus aureus | 1280 | Klebsiella sp |
| SAMN57562812 | S29 | S29 | Escherichia coli | 562 | Klebsiella sp |
| SAMN57562813 | S3 | S3 | Klebsiella pneumoniae | 573 | Klebsiella sp |

| | | | | | |
|---|---|---|---|---|---|
| SAMN57562814 | S30 | S30 | Vreelandella venusta | 44935 | Klebsiella pneumoniae |
| SAMN57562815 | S31 | S31 | Klebsiella pneumoniae | 573 | not provided |
| SAMN57562816 | S32 | S32 | Enterococcus faecalis | 1351 | Klebsiella pneumoniae |
| SAMN57562817 | S33 | S33 | Acinetobacter haemolyticus | 29430 | Klebsiella sp |
| SAMN57562818 | S34 | S34 | Pseudomonas aeruginosa | 287 | Klebsiella pneumoniae |
| SAMN57562819 | S35 | S35 | Klebsiella pneumoniae | 573 | Pseudomonas aeruginosa |
| SAMN57562820 | S36 | S36 | Pseudomonas aeruginosa | 287 | not provided |
| SAMN57562821 | S37 | S37 | Acinetobacter guillouiae | 106649 | Pseudomonas aeruginosa |
| SAMN57562822 | S38 | S38 | Acinetobacter baumannii | 470 | Pseudomonas aeruginosa |
| SAMN57562823 | S39 | S39 | Salmonella enterica subsp. enterica serovar Typhi | 90370 | Salmonella enterica serotype Typhi |
| SAMN57562824 | S4 | S4 | Klebsiella pneumoniae | 573 | Salmonella enterica serotype Typhi |
| SAMN57562825 | S40 | S40 | Candidozyma auris | 498019 | Salmonella enterica serotype Typhi |
| SAMN57562826 | S41 | S41 | Nakaseomyces glabratus | 5478 | Salmonella enterica serotype Typhi |
| SAMN57562827 | S42 | S42 | Candida tropicalis MYA-3404 | 294747 | Staphylococcus aureus MSSA |
| SAMN57562828 | S43 | S43 | Escherichia coli | 562 | Staphylococcus aureus MSSA |
| SAMN57562829 | S44 | S44 | Salmonella enterica subsp. enterica serovar Typhi | 90370 | Staphylococcus aureus MSSA |
| SAMN57562830 | S45 | S45 | Enterococcus gallinarum | 1353 | Staphylococcus aureus MRSA |
| SAMN57562831 | S46 | S46 | Staphylococcus aureus | 1280 | Staphylococcus aureus MRSA |
| SAMN57562832 | S47 | S47 | Salmonella enterica subsp. enterica serovar Typhi | 90370 | Staphylococcus aureus MSSA |
| SAMN57562833 | S48 | S48 | Streptococcus pneumoniae | 1313 | Streptococcus agalactiae |
| SAMN57562834 | S5 | S5 | Pseudomonas aeruginosa | 287 | Staphylococcus aureus MRSA |
| SAMN57562835 | S6 | S6 | Staphylococcus aureus | 1280 | Staphylococcus aureus MRSA |
| SAMN57562836 | S61 | S61 | Enterococcus faecalis | 1351 | Staphylococcus aureus MSSA |
| SAMN57562837 | S7 | S7 | Acinetobacter baumannii | 470 | Staphylococcus aureus MRSA |
| SAMN57562838 | S8 | S8 | Klebsiella pneumoniae | 573 | Streptococcus pneumoniae |
| SAMN57562839 | S9 | S9 | Enterobacter cloacae | 550 | Streptococcus pyogenes |
